# Retrospective analysis of Covid-19 hospitalization modelling scenarios which guided policy response in France

**DOI:** 10.1101/2023.12.16.23300086

**Authors:** Thomas Starck, Maxime Langevin

## Abstract

During the COVID-19 pandemic, epidemiological modelling has played a key role in public debate and policy making for anticipating the epidemic trajectory, as well as proposing and evaluating non-pharmaceuticals interventions. Despite its importance, evaluations of models’ ability to accurately represent the evolution of the disease remain scarce. Robust and systematic evaluation is needed to assess models. We investigate the following research question : were the COVID-19 scenarios proposed by modellers during the pandemic to policy-makers relevant for decision making ? To answer this, we conduct a retrospective assessment of modelling reports which guided policy response in France in 2020-2022. After systematically verifying the scenarios hypotheses (e.g., exclusion of no-lockdown scenarios when a lockdown was effectively in place), we find that out of 10 reports, reality was below the best-case scenario in 6 reports; within the best-case / worst case scenarios range in 3 reports; above the worst-case scenario in 1 report. Best-case scenarios were the closest to reality, but often came from report with a large span between best-case and worst-case scenarios beyond 2 weeks, precluding certainty about future outcomes at the time of publishing. Our results hint a systematic overestimation bias for these particular models used to anticipate epidemic evolution, which can be of importance if such models are used to contractually estimate the effectiveness of non pharmaceutical interventions. To our knowledge, this is the only national systematic retrospective assessment of COVID-19 pandemic scenarios assessing hospital burden; such an approach should be reproduced in other countries whenever possible.

**Graphical Abstract:** Reality (black line) compared to prospective scenarios (colored lines) which informed policy during the COVID-19 pandemic in France for Intensive Care Units (top) and New Hospital Admissions (bottom). Colors indicate the error between reality and scenarios, expressed as a percentage of the 1st wave peak (horizontal dashed line).

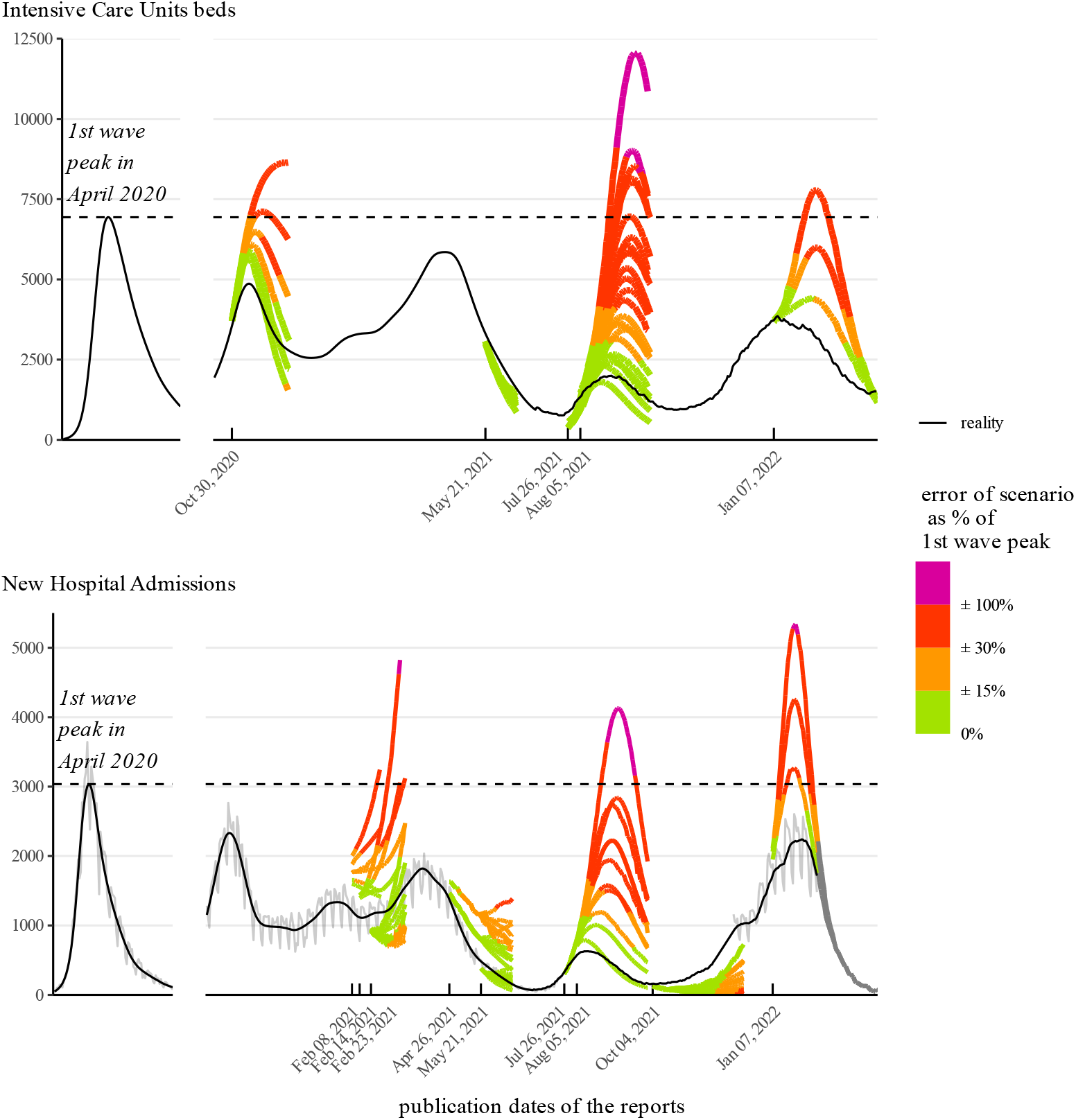

## 1 Introduction

The COVID-19 pandemic, as well as restrictions imposed to limit the spread of the virus, have had an unprecedented impact on global health, economy, and society.

Models have been central to inform decision-making, attempting to anticipate the future of the pandemic under different public health measures. Models are also used to retrospectively review the effectiveness of non pharmaceutical interventions with counterfactual scenarios [Roux et al., 2023, Flaxman et al., 2020], For instance, two thirds of the ≈ 150 studies mapped by the UK Health Security Agency to assess the effectiveness of non pharmaceutical interventions are based on modelling [UKHSA, 2023]. Models are also known to influence the societal debate through their influence on policy-makers [Sanchez, 2021].

While intended to slow virus spread, hospital burden, and mortality, the policies influenced by these models can also have harmful consequences such as increased food insecurity [FAO et al., 2023], routine child immunization disruption [Chakrabarti et al., 2023, WHO and UNICEF, 2020] and mental health issues [Léon et al., 2023], disproportionately affecting the most vulnerable groups [Li et al., 2023] and the young [UNICEF, 2021]. It is therefore essential that models correctly anticipate epidemic spread and accurately assess non pharmaceutical interventions effectiveness in order not to bias the political trade-offs: overestimating epidemic spread is not devoid of harms, as its underestimation.

In most European countries, modelling teams have been set up to inform policy making during the COVID-19 pandemic [Jit et al., 2023, Eker, 2020]. Models used include statistical models, compartmental models, meta-population models, individual-based models, and geospatial models. In the context of France, aged-structured compartmental models (such as SIR models) have been used to produce prospective scenarios [Di Domenico et al., 2021, Kiem et al., 2021] used by the French Scientific Council.

Despite its unprecedented use to support large-scale policy decisions in the COVID-19 pandemic [Eker, 2020], modelling has usually been considered to offer a relatively “low to very low” level of evidence for pandemic preparedness [WHO, 2019]. This has led to arguments for cautiousness when dealing with modelling results [Holmdahl and Buckee, 2020], as well as calls for greater model transparency, reproducibility, and validity assessments [Jin et al., 2020, Barton et al., 2020].

Epidemiological models used throughout the pandemic [Gnanvi et al., 2021], such as SIR and compartmental models [Kermack and McKendrick, 1927], are especially known to be limited in their capacity to account for local spatial heterogeneities [Zachreson et al., 2022], which can sometimes result in overestimation of disease incidence [Merler et al., 2015].

On a short time frame (typically less than 2 weeks), empirical comparison of models projections to reality shows they can be accurate [Paireau et al., 2022], but modelling scenarios informing policy-making have a longer time frame [Ferguson et al., 2020], and empirical evaluations of these latter scenarios remains scarce. Some of the available empirical analysis points towards models being unable to significantly outperform simple baselines [Chharia et al., 2022, Antulov-Fantulin and Böttcher, 2022] or failing at predicting COVID-19 outcomes [Ioannidis et al., 2022, Moein et al., 2021].

Empirical evaluations of COVID-19 modelling scenarios face several challenges. First, as explained by Nina Fefferman “in an ideal world, every epidemiological prediction of an outbreak would end up failing”, as predictions would influence policy actions that would then mitigate the outbreak and falsify the key hypotheses behind the initial model, leading to predictive failure [Jit et al., 2023, Holmdahl and Buckee, 2020]. However, honest and careful checking of models hypotheses can ensure that the comparison of models output to reality is valid, for instance by comparing models to reality only when no new new policy action have been enacted after model publication.

Furthermore, a comprehensive and systematic analysis is required in order to avoid biased cherry-picking of modelling scenarios according to whether they predicted accurately or not reality. In a related field, selective reporting of results of clinical trials has long been identified as a key source of bias. For instance, in 2000 the US Food and Drug Administration made mandatory to preregister clinical trials. This resulted in a large drop in randomized controlled trials reporting drugs benefit, from 60% to 10% [Kaplan and Irvin, 2015, Dickersin and Rennie, 2012]. Similar requirements are needed to ensure valid analysis of COVID-19 models.

Our analysis is aimed at answering the following research question: were the COVID-19 scenarios proposed by modellers during the pandemic to policy-makers relevant for decision making ?

To answer this, we focus on 3 features of the scenarios.

- **Accuracy**, which assesses whether the scenarios were close to reality.
- **Uncertainty**, which looks at the range of possibilities of the scenarios in a given report. In a most extreme case, the range could span from a negligible impact on hospital to a total submersion of the bed capacities, and would thus be uninformative.
- **Bias** of scenarios, i.e. whether modellers tend to produce outputs systematically over- or under-estimating the real epidemic transmission, inducing bias in the trade-offs the policy maker has to face.

Our case study is centered on France. We set out to perform an extensive systematic retrospective evaluation of epidemiological models that have informed policy-making in France. To ensure the relevance of our retrospective, we define clear inclusion criteria and systematically check the scenarios assumptions to confirm comparability between them with reality.

## 2 Methods

The workflow for retrieving and selecting reports for our retrospective are detailed in Figure 1 and in the following subsections (Sections 2.1 and 2.2). More details can be found in Supplementary Materials (Tables S1-S4). Since the numerical data underlying the scenarios were not public, we explain how we extracted it from the reports’ figures (Section 2.3). We finally describe our method to compare reality to scenarios and quantitatively evaluate them (Section 2.4).

**Figure 1:**
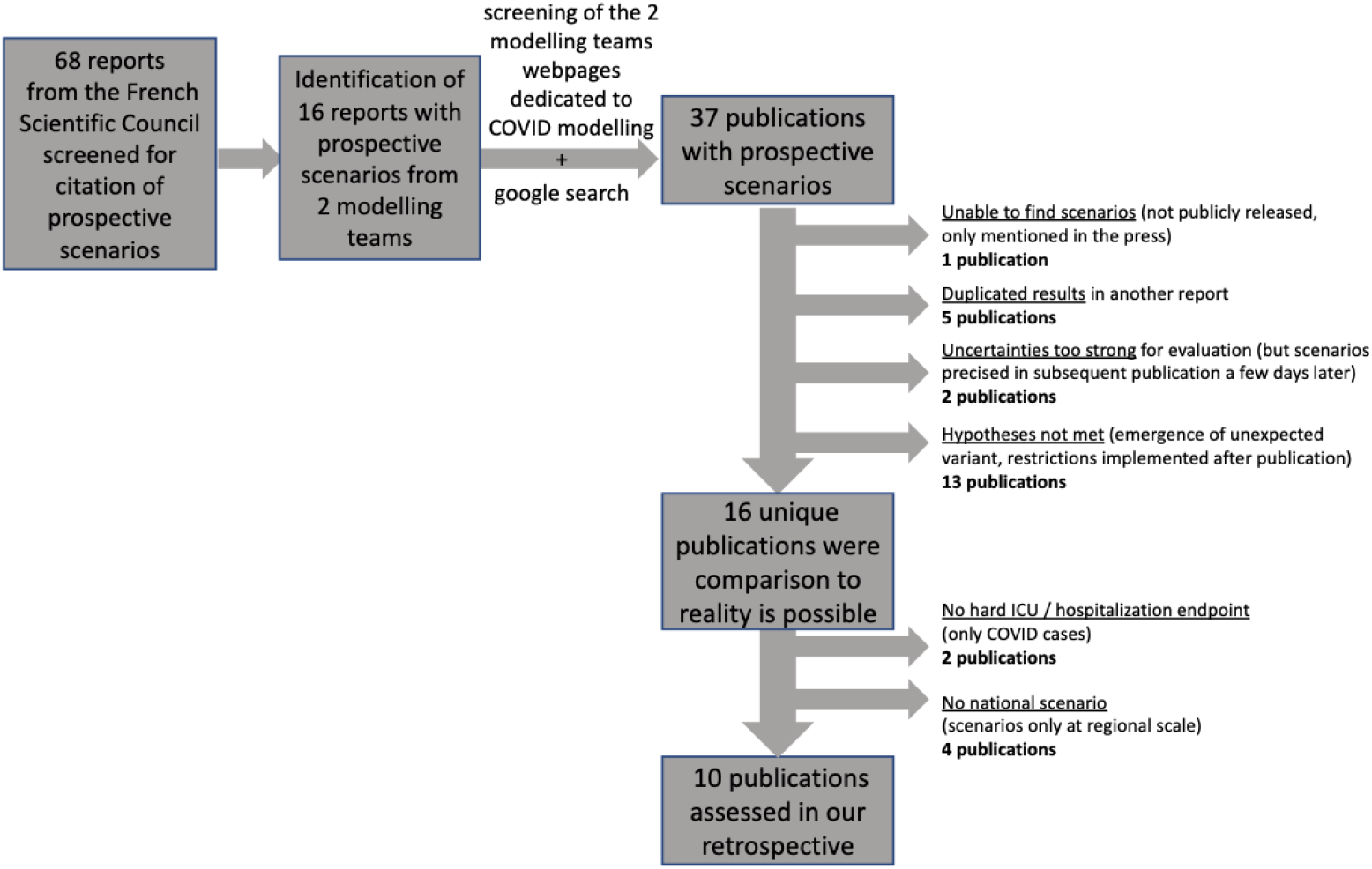
Workflow for the selection of the reports assessed in our retrospective.

### 2.1 Scope of the retrospective

To identify to modelling scenarios that potentially had a policy influence during the COVID pandemic, we first screened all the reports from the French Scientific Council, a panel of experts gathered by presidential demand at the epidemic outset [sante.gouv, 2020]. From this we identified several prospective scenarios from two French epidemiological modelling teams, the EPIcx laboratory and the Pasteur Institute modelling unit. We then went on the two units respective websites [EPIcx lab, 2023, Pasteur Institute, 2023] to check all their reports dedicated to prospectively model the COVID-19 epidemic spread. This allowed to identify other reports, not cited by the Scientific Council, but that nonetheless had media impact [Le Monde, 2021]. We also searched for mentions of scenarios from these 2 units in the media with a websearch on Google for each month from March 2020 to April 2022. The identification of all the reports is detailed in our supplementary spreadsheet and on Figure 1.

We focused on medium to long term scenarios, but excluded short-term (2 weeks) projections. We distinguished between the two types of modelling because retrospective evaluation of short-term projections is more common. For instance, Paireau et al. [2022] already self-assessed the predictive power of their short-term projections. Also, retrospective assessment of medium to long term scenarios is much less frequent because they require to check policy changes after the scenarios publications; on the contrary, short-term projections are not affected by policy changes, as their impacts manifest themselves on hospitalizations no earlier than 2 weeks after [Conseil Scientifique, 2020]. This relates to the time it takes to be infected, develop symptoms, and the condition to deteriorate sufficiently to require hospitalization.

The total number of identified reports dealing with prospective scenarios is 37. All the reports pdf and their sources are in our supplementary materials.

### 2.2 Selection and exclusion criteria

All excluded reports and the reason underlying their exclusion are featured in Supplementary Table S2 and S3 in our supplementary spreadsheet.

Of the 37 prospective reports, one in October 2020 was mentioned by the press but we were unable to recover it since the French ministry of Health did not release it publicly. Five reports feature results presented in reports already included in another report, we therefore did not evaluate them a second time. Two reports (Jan 12, 2021 and Dec 27, 2021) have too strong uncertainties in the underlying hypotheses to verify them, precluding any assessment, and are thus not analysed; however, following reports covering their scope (Feb 8, 2021 and and Jan 7, 2022) considerably reduce those uncertainties, and are included in our analysis.

To perform a fair retrospective, we only selected scenarios for which the hypotheses were verified in reality. For instance, we would discard a scenario which assumes a “no lockdown” situation, but where a lockdown was put in place shortly after the publication of the report. This is done for the different major non pharmaceutical interventions that could impact the modeled endpoints, such as lockdowns or curfews, but also for emergence of a new variant, or for vaccination rates. This alleviates a common form of circular reasoning which claims that epidemiological models were unaccurate precisely because they lead to measures that changed the underlying hypotheses. All details concerning our hypotheses verification are reported in Tables S3 and S4, and particular examples are given in Table 1. Verifying these hypotheses further excluded 13 publications.

**Table 1:** Example of hypotheses verification for scenarios inclusion or exclusion in our retrospective analysis.

| Report Date | Included ? | Justification |
| --- | --- | --- |
| Oct 26, 2020 | no | national lockdown implemented 2 days later not considered in scenarios |
| Oct 30, 2020 | yes | scenarios consider national lockdown announced 2 days earlier |
| Jul 9, 2021 | no | implementation of health pass announced 3 days after |
| Jul 26, 2021 | yes | health pass implementation considered through estimations of R decrease |

This yields 16 reports, whose scenarios endpoints are reported in Table 2. We restrict our retrospective study to reports modelling strong endpoints, i.e. hospitalizations and Intensive Care Units. Deaths would be another strong endpoint but is never modelled in the reports. We discard positive cases as this endpoint depends heavily on the testing rate which varies through time depending on people’s behavior, and is less important for decision-making than Intensive Care Units (ICU) and hospital caseloads. This criteria further excluded 2 publications (Feb 21, 2022 and Mar 10, 2022).

**Table 2:** Scenarios endpoints reported in the 16 reports eligible for comparison to reality after hypotheses validation. We did not analyze the last 2 reports only focusing on positive Covid-19 cases as our review focuses on hospital and ICU hard endpoints (see text). We also did not analyze the 4 reports which did not report national data. The total number of reports analyzed in our study is thus 10.

| Report Date | ICU beds | ICU admissions | Hospital beds | Hospital admissions | Positive Cases | National scenarios | Regional scenarios |
| --- | --- | --- | --- | --- | --- | --- | --- |
| Apr 12, 2020 | x | x |  |  | x |  | x |
| Apr 28, 2020 | x |  | x |  |  |  | x |
| May 12, 2020 | x | x |  |  | x |  | x |
| Oct 30, 2020 | x | x |  |  |  | x |  |
| Nov 8, 2020 |  |  |  | x |  |  | x |
| Feb 8, 2021 |  |  |  | x | x | x |  |
| Feb 14, 2021 |  |  |  | x |  | x | x |
| Feb 23, 2021 |  |  |  | x | x | x |  |
| Apr 26, 2021 |  |  |  | x |  | x |  |
| May 21, 2021 | x |  | x | x |  | x |  |
| Jul 26, 2021 | x | x |  | x | x | x |  |
| Aug 5, 2021 | x | x | x | x |  | x | x |
| Oct 4, 2021 |  |  |  | x |  | x |  |
| Jan 7, 2022 | x |  | x | x |  | x |  |
| Feb 21, 2022 |  |  |  |  | x | x |  |
| Mar 10, 2022 |  |  |  |  | x | x |  |
| Total | 8 | 5 | 4 | 10 | 7 | 12 | 6 |

We also only looked at publications reporting national data, excluding reports only reporting regional scenarios (4 reports). This allows a common comparison between the different reports, as the studied regions are not always the same. Also, including regional scenarios would have multiplied the number of analysis and given too much weight to some particular periods during the pandemic: for instance, the Nov 8, 2020 reports studies 5 different regions, and the Aug 5, 2021 report 12 regions.

Our final analysis thus focus on 10 different reports, of which only one (Feb 14, 2021) is from the EPIcx lab. Among the remaining 10 reports, we focus on the most commonly reported endpoints, i.e. Intensive Care Units beds occupancy and hospital admissions (Table 2).

### 2.3 Data extraction and preparation

None of the reports provided their scenarios data in open data. We manually extracted the data from the original figures, using WebPlotDigitizer https://automeris.io/WebPlotDigitizer/.

For each report, we first excluded the scenarios where hypotheses were not met, and then extract the remaining scenarios of interest. We also extracted the reality data available up to the report publication date (see Figure 2). This allows us to check if our manual extraction was carried out correctly, by comparing the reports reality data to French official hospitalization and Intensive Care Units data.

**Figure 2:**
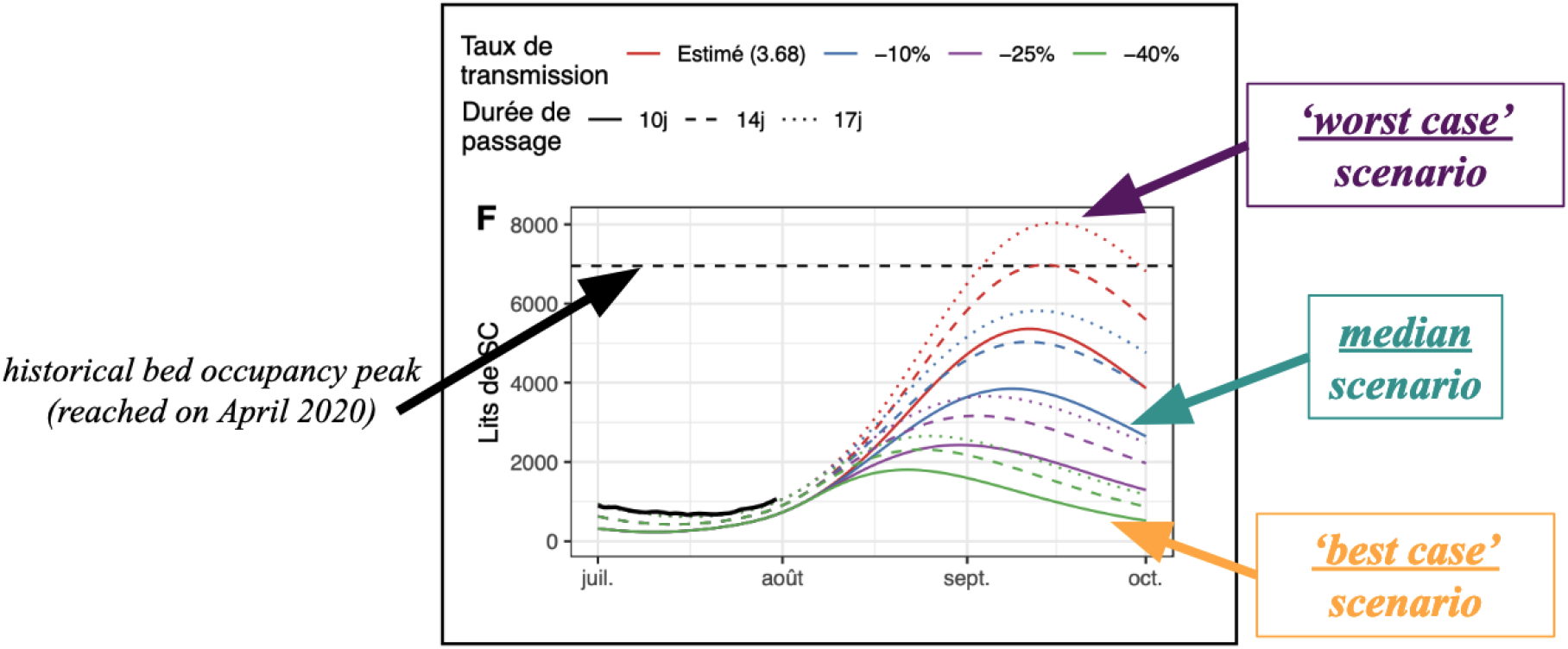
Screenshot example of the original scenarios published in the Aug 5^*th*^, 2021 report. Some of the scenarios hypotheses are detailed in the top legend. Black solid line: real data available at publication date and used by modelers for calibration. Colored lines: scenarios. We indicate how we define the median, best-case and worst-case scenarios for our study. We use the historical peak (horizontal dashed line) to normalize the scenarios error.

Most of our official reality data comes from Paireau et al. [2022], but this source stops on July 2021. This is the most reliable source for scenarios comparison to reality, since it comes directly from one modelling team, and includes their own pre-processing and cleaning procedure (see Paireau et al. [2022] for details). For the rest of the period, we use either French official data data.gouv [2023] or reality data manually extracted from the reports.

### 2.4 Evaluating the scenarios

As illustrated in Figure 2, each report provides multiple scenarios. While it is likely that some scenarios were favored as more probable during interactions between modelers and policy-makers, this information is not available. Our retrospective evaluation focuses on 3 particular scenarios in each report: the worst-case, the median and the best-case scenarios.

We use standard quantitative metrics to asses scenarios uncertainty (section 2.4.1), accuracy (section 2.4.2) and bias (section 2.4.3). Each metric is computed over the whole report period (starting at date of publication), and by 2-week periods to see their evolution through time.

#### 2.4.1 Uncertainty

As multiple scenarios are proposed, a key aspect is the uncertainty (or conversely, the informativeness) of a report. We define the uncertainty as the difference between the values anticipated by the worst-case and best-case scenarios (see Figure 2). For each report, we compute the average uncertainty over the considered time period.

#### 2.4.2 Accuracy

We assess accuracy for 3 representative individual scenarios in each report: the worst-case scenarios (with the highest anticipated values), the best-case scenario (with the lowest anticipated values), as well as the median scenario (see Figure 2).

For each of these scenarios, we assess their accuracy with the Mean Absolute Error (MAE) and the Mean Absolute Percentage Error (MAPE).

- The Mean Absolute Error 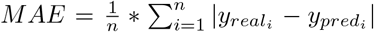 is a widely used metric for evaluating the accuracy of models. It measures the average absolute difference between the predicted values and the real values, where 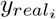 is the actual value at the time step *i*, 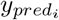 is the predicted value, and n is the number of time steps. In our case, it is expressed in number of beds per day.
- The Mean Absolute Percentage Error 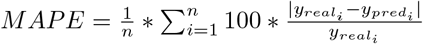 is similar to Mean Absolute Error but is normalized with respect to the real values. It measures the average absolute difference between the predicted values and the real values divided by the real values, where 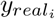 is the actual value at the time step *i*, 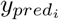 is the predicted value, and n is the number of time steps. Unlike the Mean Absolute Error, it is expressed in percentage of the real value and is comparable across different endpoints (i.e. Intensive Care Units beds and new hospitalizations).

#### 2.4.3 Bias

Previous accuracy metrics express the error in absolute terms, but do not indicate the direction of the error.

To address this limitation, we use the mean error. It indicates whether on average the scenario tended to overestimate (values >0) or to underestimate (values <0) the reality, and can thus be used to identify systematic errors in a forecast.

- The Mean Error 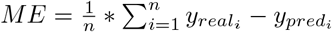 is a metric that measures the average difference between the predicted values and the real values. The Mean Error is positive when the scenario overestimates the reality on average and negative when it underestimates the reality on average. In our case, it is expressed in number of beds per day.

#### 2.4.4 Normalization by historical peak

All the above metrics except Mean Absolute Percentage Error are expressed in terms of number of Intensive Care Units beds or new hospitalizations per day. We normalize these metrics by comparing them to the maximum historical values reached by these two endpoints during the pandemic, based on Paireau et al. [2022] data : 6937 for Intensive Care Units beds occupancy and 3036 for (smoothed) daily new hospitalizations. These peaks can be visualized on Figure 3.

**Figure 3:**
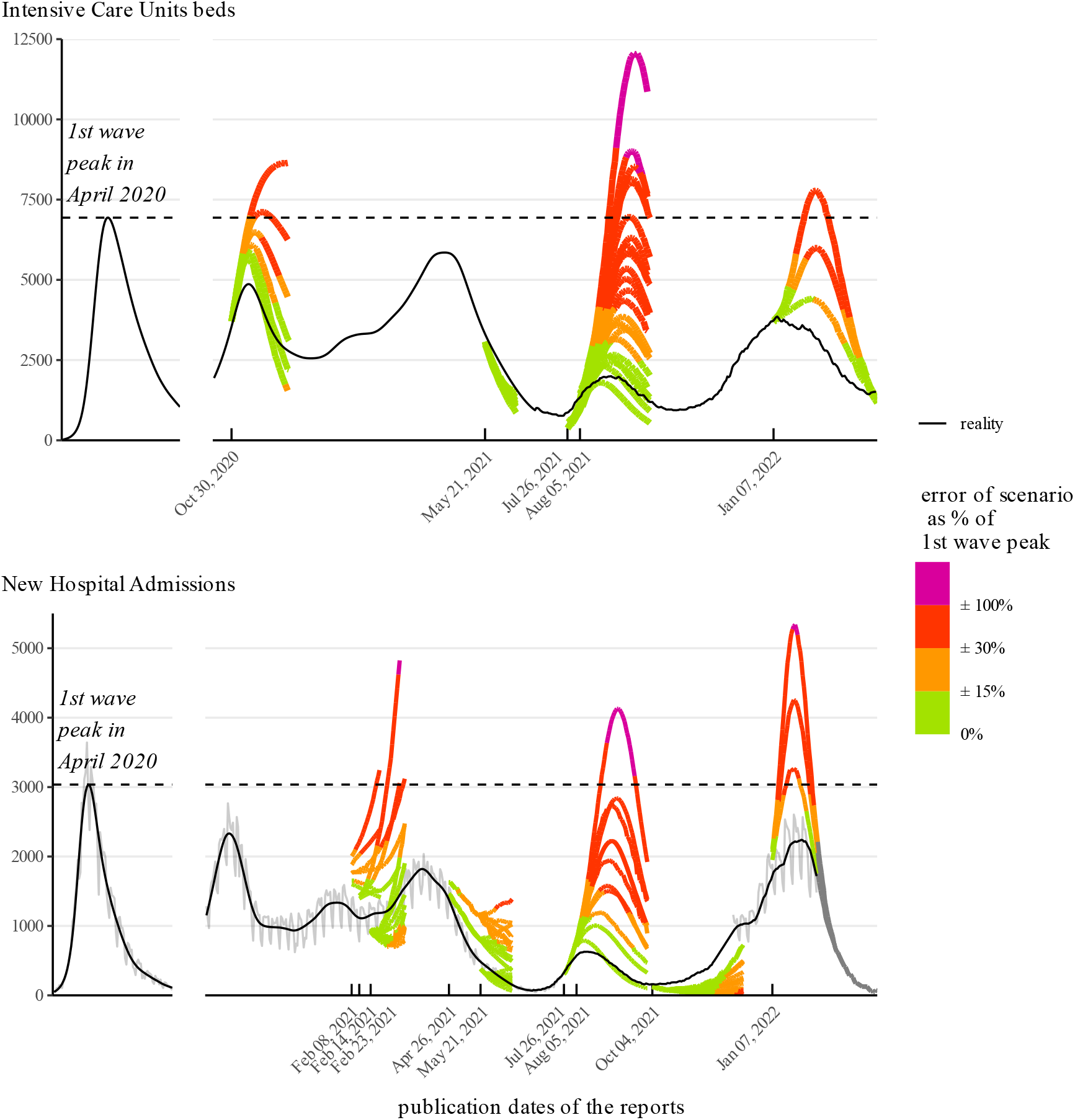
Reality (black line) compared to prospective scenarios (colored lines) which informed policy during the COVID-19 pandemic in France for Intensive Care Units (top) and New Hospital Admissions (bottom). Colors indicate the error between reality and scenarios, expressed as a percentage of the 1st wave peak (horizontal dashed line). Note that an error of *±*15% (green) means a possible range of values of 30% of the historical peak.

Normalizing allows to express metrics in a scale that is relevant for policy-making, as a percentage of historical peak, and to compare scenarios with different endpoints (Intensive Care Units and new hospitalizations). Also, a drawback of the relative MAPE is that when hospitalizations are low, small absolute errors can result in high relative errors; normalization of absolute error avoid this pitfall.

All metrics are summarized in Table 3.

**Table 3:** Summary of metrics used for evaluating scenarios.

| Assessment Goal | Metric | Computed over | Interpretation |
| --- | --- | --- | --- |
| Uncertainty | Average Uncertainty | all scenarios from one report | What is the possible span of values according to the scenarios |
| Accuracy | Mean Absolute Error (MAE) | one specific scenario (e.g. best-case, median or worst-case scenario) | Average error between the scenario and the reality |
|  | Mean Absolute Percentage Error (MAPE) | one specific scenario | Average error between the scenario and the reality, with the error expressed as a percentage of real values |
| Bias | Mean Error (ME) | one specific scenario | Whether and how much the scenario, on average, overestimated or underestimated reality |

## 3 Results

### 3.1 Qualitative comparison of scenarios to reality

#### 3.1.1 Intensive Care Units beds

Figure 3 (top) compares reality to the scenarios of the 5 reports anticipating Intensive Care Units beds occupancy.

In 3 reports (Oct 30, 2020; Jul 26, 2021 and Jan 07, 2022) reality was below the best-case scenario. For the Aug 5, 2021 report, reality corresponded to the best-case scenarios. One report (May 21, 2021) has a median scenario close to reality. This indicates a general bias towards scenarios more pessimistic than reality. These latter 2 more accurate scenarios (May 21 and Aug 5, 2021) are updates of previous reports (Apr 26 and Jul 26, 2021).

The May 21, 2021 report is also the only one with a low span between the best-case and the worst case scenarios. Two of the reports (Jul 26, 2021; Aug 5, 2021) feature scenarios whose Intensive Care Units maximum occupancy range from close low occupancy to higher than the 1st wave peak, providing little certainty. The scenarios from the two remaining reports (Oct 30, 2020 and Jan 07, 2022) have a span of about 50% of the historical peak.

#### 3.1.2 Hospital Admissions

Figure 3b compares reality to the scenarios of the 9 reports anticipating hospital admissions.

Out of these 9 reports, 4 have reality below their best-case scenario (Feb 14, 2021; Apr 26, 2021; Jul 26, 2021; Jan 7, 2022) and 2 have reality reaching their best-case scenario (Feb 8, 2021 : Aug 5, 2021). For 1 report (Oct 4, 2021), reality is above the worst-case scenario. This leaves 2 reports (Feb 23, 2022; May 21, 2021) where reality is within the range of featured scenarios, between the best and worst case.

Thus, about half (5/9) of the reports on hospital admissions have a scenarios range outside the best/worst case range. Moreover, two thirds (6/9) of reports have their best-case scenario equal or above reality, which points to a bias towards scenarios more pessimistic than reality.

#### 3.1.3 Short-term change of the results

In 3 instances (Feb 2021; Apr-May 2021; Jul-Aug 2021) several reports are published within a short time period, about 1 month or less. This allows to visualize how sensitive the modelers’ output is to the epidemic short-term dynamic (figure 4).

**Figure 4:**
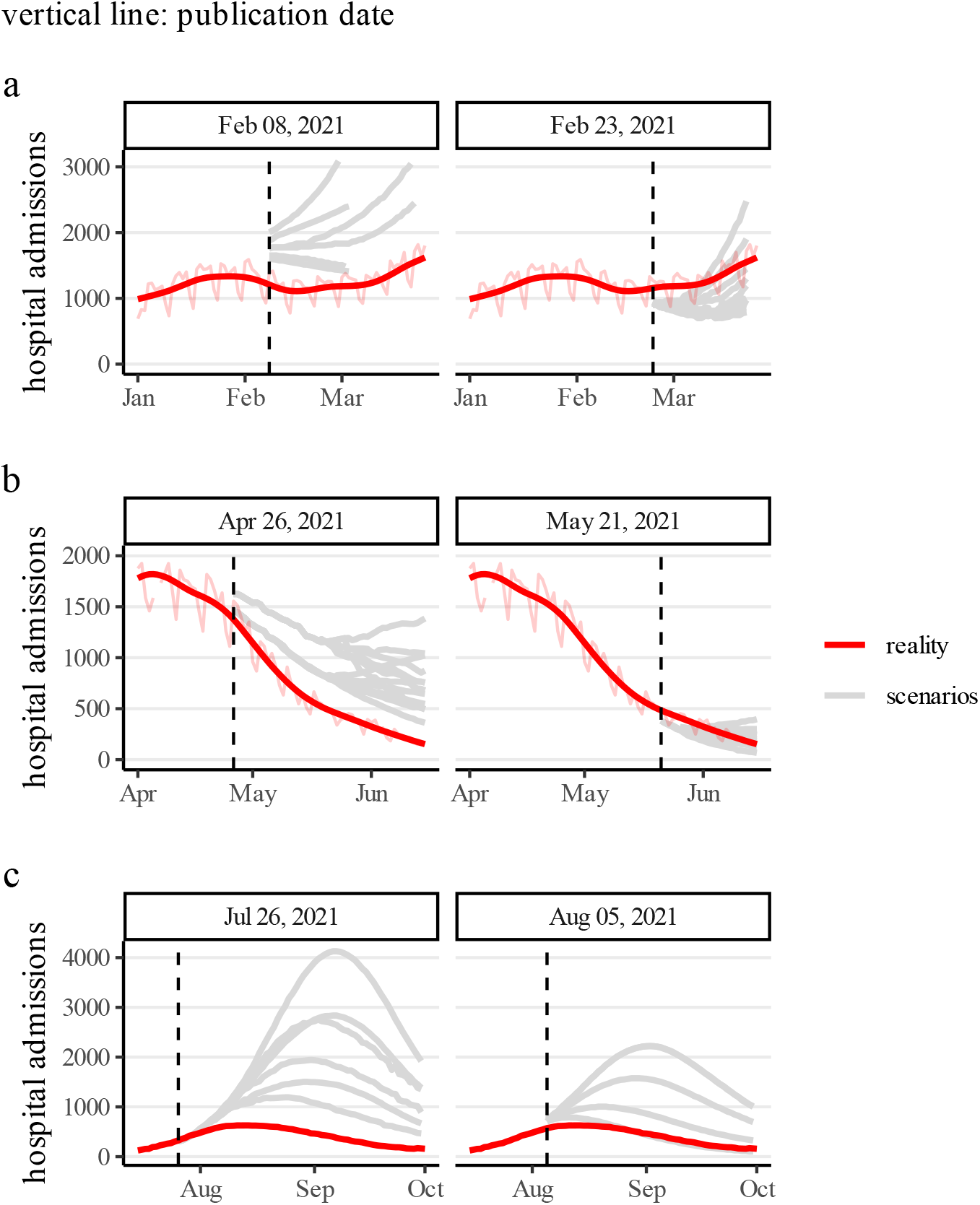
Comparison of scenarios variations for reports published a few weeks apart. **a)** scenarios during winter 2021. **b)** scenarios during spring 2021 **c)** scenarios during summer 2021.

For the Feb 2021 period (Figure 4a), in the first Feb 8 report, a sensitivity analysis performed by the modelers suggests the possibility of both a small decline and an exponential increase, and reality is slightly lower than the best-case scenario. In the final re-assessment of Feb 23, the reality falls within the scenarios range.

For the Apr-May 2021 period (Figure 4b), the first report (Apr 26) presents a range of scenarios which, after a first decline, features dynamics ranging from downwards to stagnating trends. The final range of these scenarios represent one third of the historical 1st wave peak. Reality was eventually below the best-case scenario. The report update (May 21) features scenarios which are below the best-case scenario of the original report. Reality falls within the scenarios range of this updated report.

Finally, for Jul-Aug 2021 (Figure 4c), 2 reports were published within 10 days. While the historical 1st wave peak was about 3000 daily hospital admissions, the Jul 26 scenarios ranged from 1000 to 4000; reality was half the best-case scenario. The Aug 5 update presents scenarios about 2 times smaller than the original report, and reality corresponds to the best-case scenario.

In all 3 instances, results are updated towards more optimistic scenarios compared to the first publications. These 3 instances correspond to the 3 reports where reality falls between the best case and worst case scenarios. In the 7 remaining original reports, reality is outside the scenarios range, one time above the worst case scenario (Oct 4, 2021), the six other times below the best-case scenario.

### 3.2 Quantitative performance assessment

In this section we present a systematic analysis of the modelling reports using the quantitative metrics presented in Table 3.

#### 3.2.1 Uncertainty

Figure 5 shows the distribution of scenarios average uncertainty for each published report, by 2-weeks periods. Uncertainty is usually low (<20% of the historical peak) during the first 2-weeks period, but logically increases through time.

**Figure 5:**
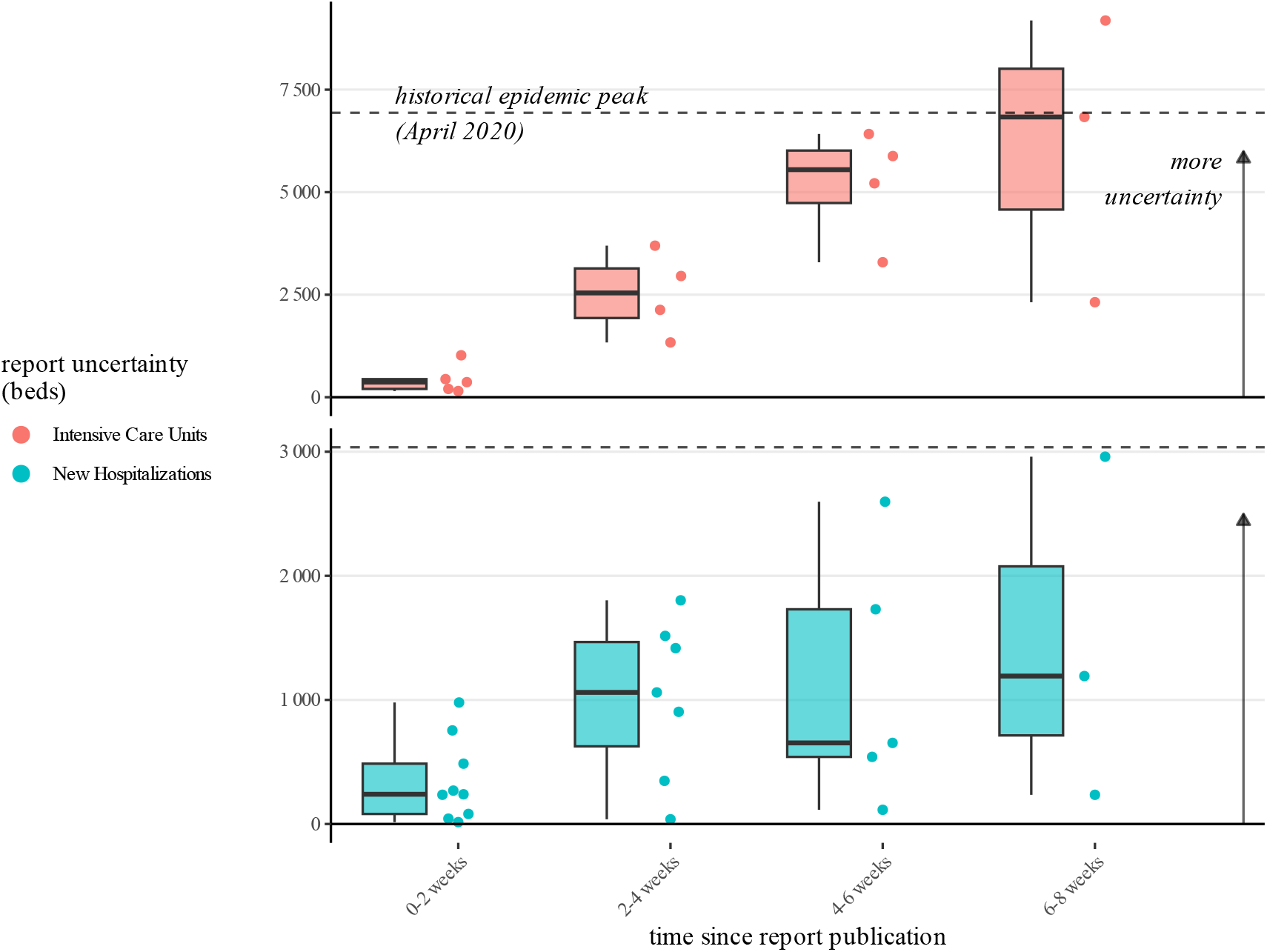
Average uncertainty of scenarios range in the modelling reports, for each 2-week period since report publication. Top: Intensive Care Units. Bottom: New Hospitalizations.

For Intensive Care Units beds, the median of the average uncertainty across reports is around 2500 beds at 2-4 weeks (compared to an historical peak of about 7000 beds, i.e. an uncertainty of about 30% of the peak). After one month, the median uncertainty is close to the historical peak (see for instance July 26 and Aug 5 scenarios in Figure 3 top).

After 2 weeks, median uncertainty related to scenarios focusing on hospital admissions is about one third of the epidemic peak and stays around this value, but with large differences between reports. At one month, uncertainty spans from close to zero to the historical peak.

#### 3.2.2 Accuracy

Figure 6 shows the Mean Absolute Error of the median, worst-case and best-case scenario of each report. A complementary measure is presented in 8b as the Mean Absolute Percentage Error.

**Figure 6:**
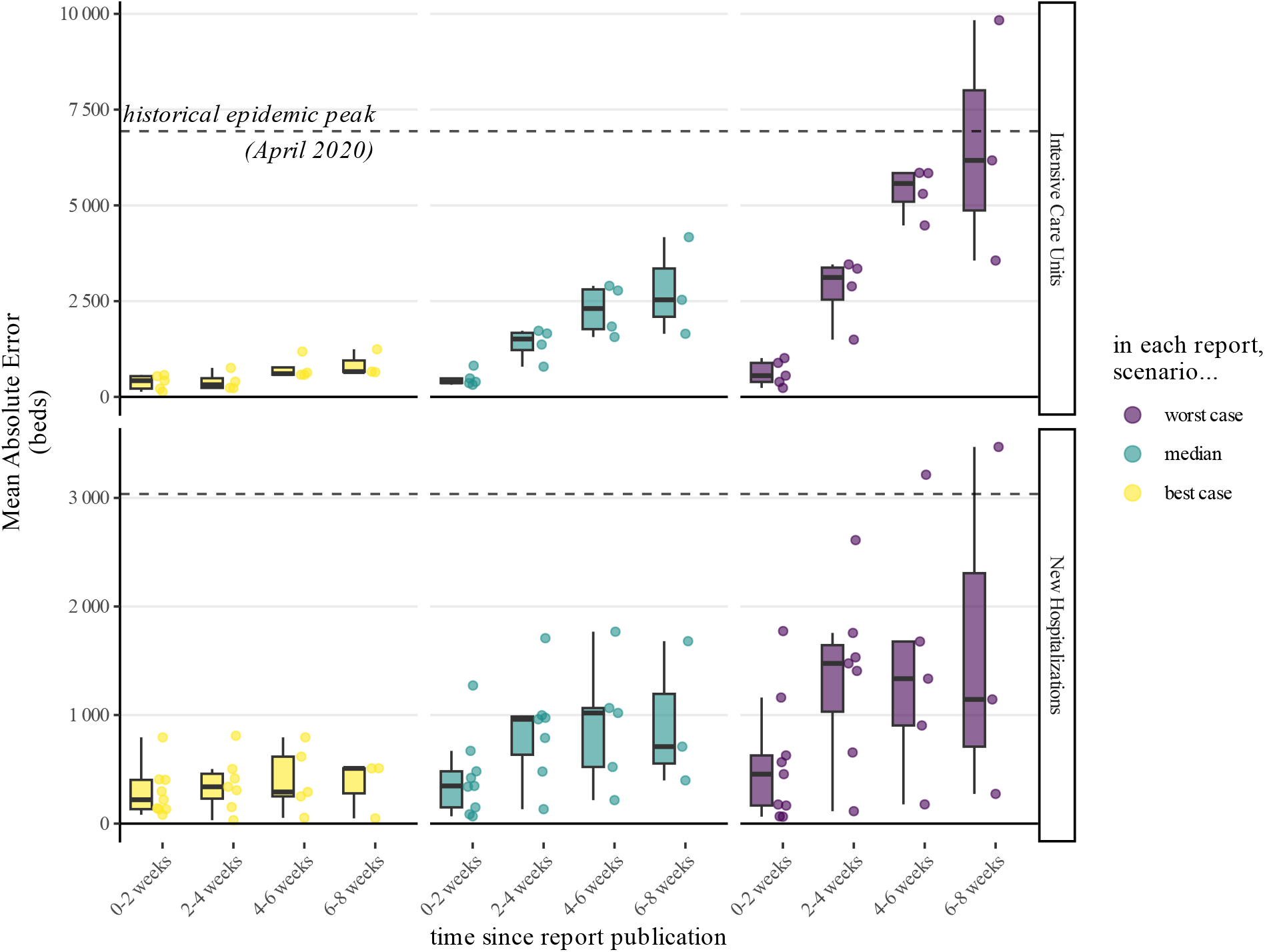
Accuracy of the modelling reports for the worst-case scenarios (purple), median scenarios (blue) and best-case scenarios (yellow) for intensive care units (top) and new hospitalizations (bottom). The best-case scenarios are the most accurate.

We remind that a scenario with e.g. ±20% mean absolute error means that its deviation from reality is ±20% of the epidemic peak, i.e. an interval of 40% the epidemic peak.

During the first 2 weeks, errors of best-case, median and worst case scenarios are similar, owing to the low uncertainty of the reports during this time frame (see previous section).

After this 2-weeks period, for worst-case and median scenarios, the mean absolute error increases, but remains about the same for best-case scenarios (Figure 6). For these best-case scenarios, the median MAE mostly stays close or below 15% of the epidemic peak.

After one month, the median MAE of the median scenarios is about one third of the epidemic peak, with individual scenarios spanning from close to 0 to two thirds of the epidemic peak. For worst-case scenarios, the median is around half the epidemic peak for New Hospitalizations and close to the epidemic peak for Intensive Care Units beds occupancy.

These results must be further context contextualized with the uncertainty of the reports. Producing numerous scenarios with a large span guarantees one will be close to reality, but would be of little use. That is why we analyse both uncertainty and accuracy together in the following section.

#### 3.2.3 Uncertainty vs Accuracy

The scenarios from Jul 26 2021 in Figure 3 exemplify the dichotomy between accuracy and uncertainty. The report scenarios go from a peak smaller than any seen since the start of the epidemic to way higher than the first wave peak. Reality finally came close to the best case scenario, but had the epidemic followed a catastrophic trajectory, the worst-case scenarios would have been close to reality. Producing many different scenarios ensures at least one will be accurate.

Figure 7 synthesizes this accuracy/uncertainty dichotomy across all reports. For each 2-weeks period since the publication of a report, it compares the accuracy of the scenarios (vertical axis) to the uncertainty of the report they are issued from (horizontal axis).

**Figure 7:**
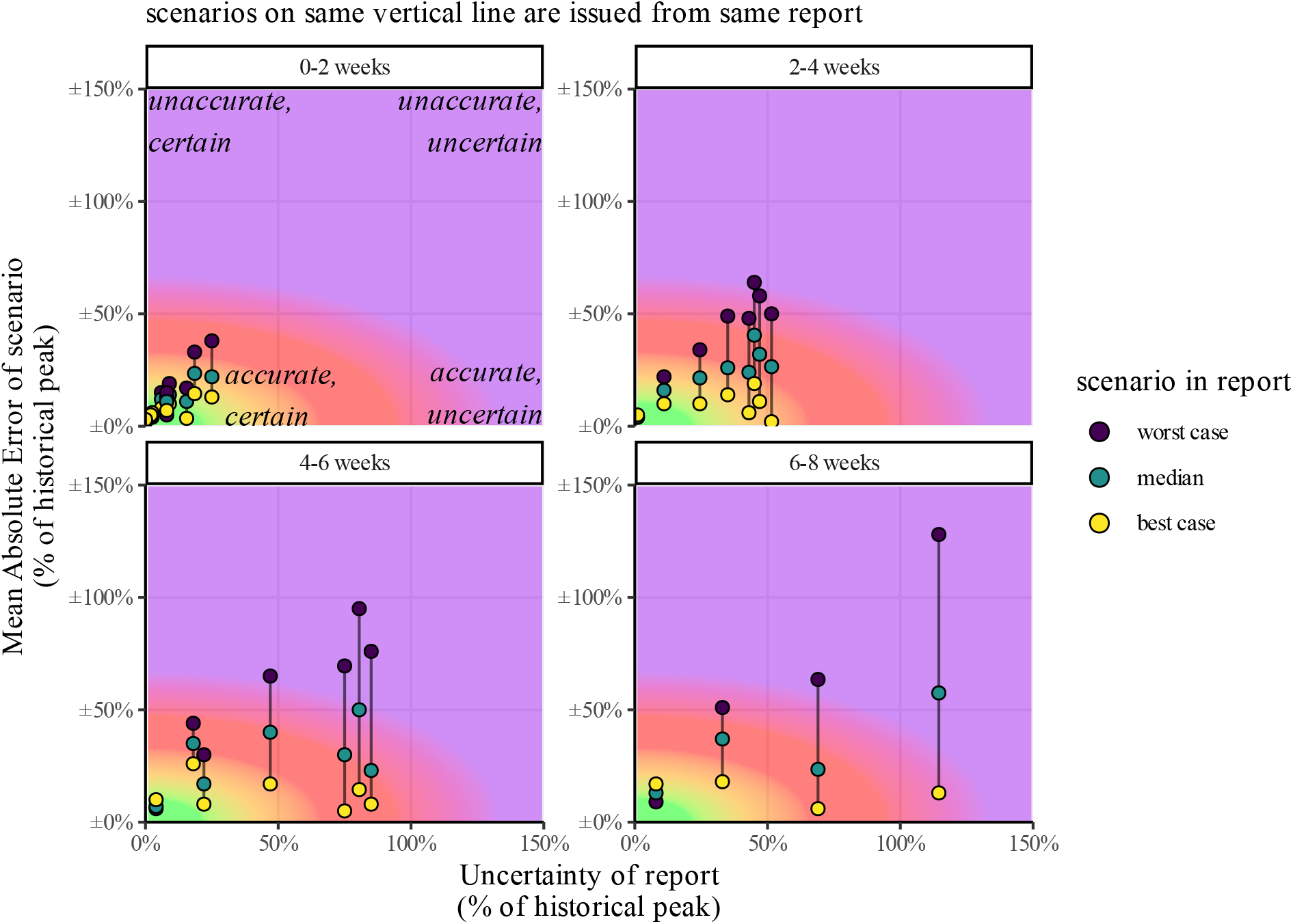
Uncertainty and Accuracy of scenarios. For each scenario (point), and for each 2-weeks period since report publication (panes), x-axis represents the mean uncertainty of the report and y-axis represent the Mean Absolute Error of the scenario. All values are expressed as percentage of historical peak. Scenarios on a same vertical line are issued from the same report. Note that an error of *±*50% means a confidence interval of 100% of the historical peak: predicting a value of 50% the historical peak with *±*50% error means reality can be anywhere between 0% and 100%.

We define a scenario as accurate if its Mean Absolute Error is 15% of the historical peak. As the absolute error can relate to both under or over estimation, this means that the range of accepted values around reality for our “accurate” definition is 2 *∗* 15% = 30% of the historical peak.

We define a report as certain if its uncertainty is below 30% of the historical peak. The combination of these 2 criteria is represented in green on Figure 7.

While most accurate scenarios come from a report with low uncertainty during the first 2 weeks, there numbers falls to 3 out of 8 at 2-4 weeks, 2 out of 6 at 4-6 weeks, and 1 out of 4 at 6-8 weeks.

#### 3.2.4 Bias

Previous error metrics focus on absolute errors, which does not indicate whether scenarios under or overestimate reality. From visual inspection (Figures 3), we see that the majority of reports overestimate the real epidemic activity, and that the errors discussed above corresponded to overestimation. To quantitatively evaluate this, we compute the Mean Error for median, best-case and worst-case scenarios.

In Figure 8a, the distribution of Mean Error across all reports is presented for the best-case, median, and worst-case scenarios. Unbiased reports, which do not consistently overestimate or underestimate the modeled endpoints, would exhibit a distribution of mean errors centered around zero for the median scenarios, while best-case and worst-case scenarios would respectively be centered around negative and positive values.

**Figure 8:**
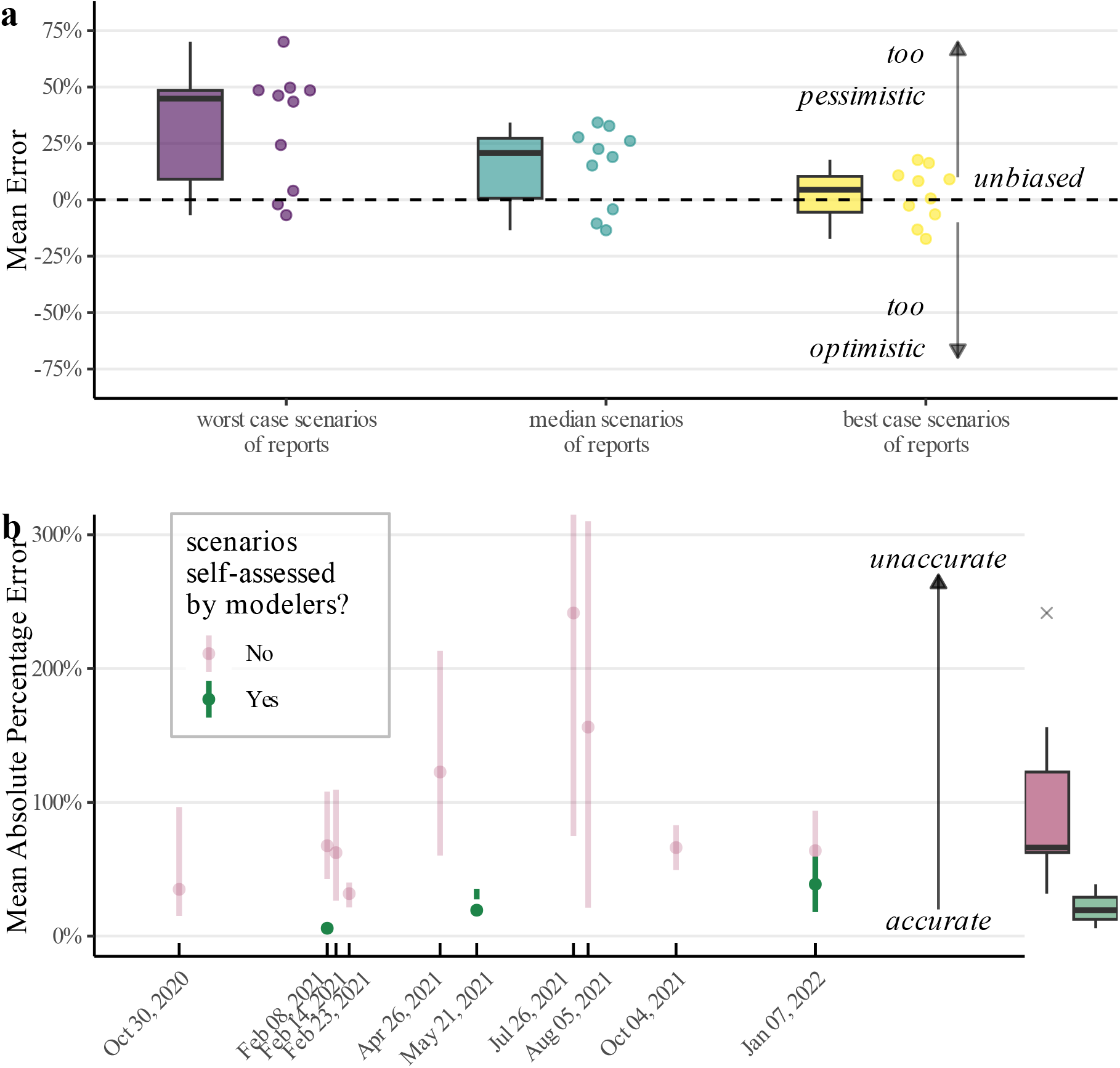
**a)** bias of best-case (yellow), median (blue) and worst-case (purple) scenarios of each report, assessed with the Mean Error. Compared to the real epidemic activity, values close to 0% are unbiased, negative values are too optimistic and positive values are too pessimistic. The best-case scenarios are unbiased, while median and worst-case scenarios are more pessimistic than reality. **b)** for each report, accuracy of scenarios (assessed by mean absolute error) whether they are publicly self-assessed by modelers (green) or not (red). Boxplots on the right refers to the distribution of median scenarios across all reports. Reports self-assessed by modelers are more accurate than the complete distribution.

In this retrospective case, the median scenarios displays a bias towards overestimation. To have an unbiased assessment, one would have to focus on the reports best-case scenarios.

#### 3.2.5 Modelers’ own self-assessment

Out of the 16 reports where comparison to reality is appropriate after hypotheses verification (Table 2), we find 4 instances where the modelers performed a public retrospective assessment (Figure 8b), by directly plotting reality against their scenarios, as we did in this article. One of the instances concerns a report not included in our retrospective as it only focuses on COVID cases (Feb 21, 2022 report, see Methods Section).

For the remaining 3 reports, one is identical to our own comparison (May 21, 2021, Cauchemez [2021]); one does not discuss the hypothesis related to vaccine efficacy, leading to a slightly different comparison than ours (Jan 7, 2022 report, see Figure 9c and d); the last one features scenarios that are not present in the original report (Feb 8, 2021 report, see (Figure 9a and b).

**Figure 9:**
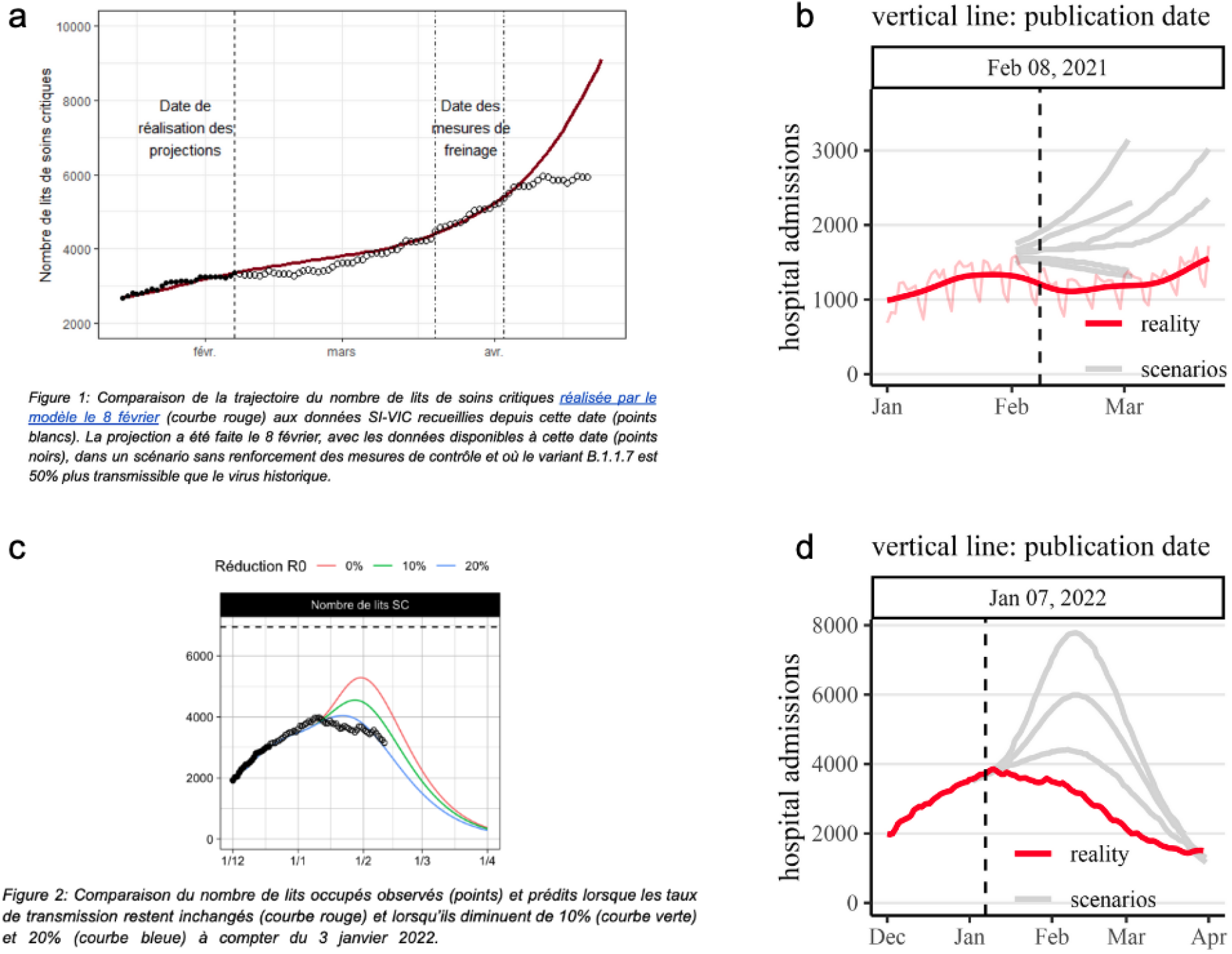
Self-assessments by modelers which are different than ours. Top: for February 8, 2021 report. Bottom: for Jan 7, 2022 report. **a)** self-assessment made by modelers, based on a curve absent from the original report. **b)** reality vs scenarios actually present in the original report. **c)** self-assessment made by modelers, based on the subset of scenarios with high vaccine efficacy assumptions (see text and supplementary materials). **d)** reality vs scenarios with low vaccine efficacy assumptions.

Concerning the Feb 8, 2021 self-assessment (performed in the Apr 26, 2021 report), the comparison displayed by the modelers can be see on Figure 9a. However, the curve presented does not correspond to any published scenario in the actual Feb 8 report. We find no mention of the actually published scenarios (featured on Figure 9b) in the modelers’ self-assessment.

Concerning the Jan 7, 2022 report (performed in a Feb 15, 2022 report), the modelers originally presented 2 sets of scenarios regarding vaccine efficacy against the Omicron variant. But in the retrospective assessment, only the optimistic scenarios are displayed (Figure 9c). We estimate that, based on data from UK Health Security Agency available at that time UKHSA [2023], the pessimistic assumptions are more correct (see discussion in Supplementary Materials). While there is obviously room for debate concerning the most correct set of scenarios, there is no discussion on the vaccine efficacy hypothesis in the modeler’s self-assessment, so we cannot compare their justification to ours.

Figure8b compares the accuracy of the scenarios in all the reports of our retrospective and in the subset of reports self-assessed by the modelers. Reports self-assessed by the modelers have a lower Mean Absolute Percentage Error than the ones they did not assessed. A more visual and qualitative comparison is available in the supplementary materials (Figure S1).

## 4 Discussion

### 4.1 Focus on worst-case scenarios

While this retrospective deals simultaneously with worst-case, best-case and median scenarios, it is often worst-case scenarios which play the major role in policy-making and public debate. For instance, the worst-case scenario from the October 26th, 2020 report was described as “unavoidable” by the head of the French government when announcing a national lockdown Macron [2020].

Yet, in our retrospective, worst-case scenarios appear to be the most unaccurate. For Intensive Care Units beds, the median error at one month is close to 6000 beds, considering that the historical peak was around 7000 beds (Figure 6). On the other hand, best-case scenarios were less discussed by policy-makers during the pandemic, but were on average more accurate than median and worst-case scenarios (Figure 6, Figure 8a).

### 4.2 Common critics to scenarios evaluation

#### 4.2.1 Scenarios trigger social distancing measures, resulting in lower epidemic spread

A common critic of retrospective analyses of modelling scenarios is that models lead to policy change, avoiding the projections made by the models Jit et al. [2023], Holmdahl and Buckee [2020].

This is why we stress the fact that we only assess scenarios in which the underlying hypotheses were met in reality, and exclude other scenarios. If a policy change (e.g. curfew or national lockdown) which was not explicitly modeled takes place after publication, we would not run our retrospective on this specific scenario. After this exclusion, the remaining scenarios variability in a given report lies in different effective reproduction number *R*, displaying irreducible uncertainty in the effectiveness of the non pharmaceutical interventions.

Careful checking of all modelling hypotheses, as well as detailed justification of every included and excluded scenario, is given in the supplementary materials.

#### 4.2.2 Better safe than sorry

Additionally, it may be argued it is better to anticipate worst-case scenarios event at the expense of overestimating future epidemic spread, because it allows to take measure avoiding the most dire consequences.

However, non pharmaceutical interventions like lockdown or school closure aimed at slowing epidemic spread can be associated with adverse harms, including rise in food insecurity FAO et al. [2023], medical care and routine child immunization disruption Chakrabarti et al. [2023], WHO and UNICEF [2020], rise in mental health issues Léon et al. [2023] and economic disruption.

If overestimation is accepted for modelling epidemic outcomes, there is no reason that other modelling studies trying to anticipate adverse consequences of social distancing measures could not themselves aim at being “better safe than sorry”, e.g. by drastically overestimating the rise in food insecurity. But one would then simply find themselves with multiple overly pessimistic studies pointing in opposite trends, not facilitating decision.

Therefore, accuracy is critical for modelling aimed at informing policy-makers and citizens who have to arbitrate trade-offs on multiple dimensions.

### 4.3 Consequences for Non Pharmaceutical Interventions evaluation

Some models are used to assess the effectiveness of non pharmaceutical interventions, by providing counterfactual scenarios of what would have happened in the absence of the implemented measures, e.g. a lockdown [Roux et al., 2020]. The difference between this counterfactual and reality is then deducted as the effect of the non pharmaceutical intervention.

However, if a model has a systematic bias towards overestimating epidemic activity, comparing the reality to the modeled alternative without intervention leads to overestimating the effect size of the intervention.

Our retrospective might hint at a potential systematic overestimation bias in the particular models used to inform policy-making in France during the COVID-19 pandemic. This is consistent with studies reporting that the homogeneity hypothesis in compartmental SIR models can lead to inflating the epidemic spread Merler et al. [2015], Zachreson et al. [2022].

Another possibility is that models themselves are accurate, but the remaining hypotheses made regarding the effectiveness of the different interventions (translated in different reductions of the reproduction number R in the reports) are systematically too pessimistic, underestimating their effectiveness.

Unfortunately, it is difficult to chose between these 2 possibilities, since the reproduction number is both a model feature and influenced by the model hypotheses.

To illustrate this, let’s take the self-assessment report regarding the Omicron wave (Feb 15, 2022 report), where modelers observe that *“the observed trajectory is close to the scenario in which it was assumed that the French would reduce their contacts by* 20% *in January”*. Falsifying the 20% contact reduction hypothesis would require empirically measuring contact in the French population, which is difficult. If anything, google mobility data indicates no major change in France during the period (Jan 7 to Feb 15, 2022) Google [2022], but even this is only an approximate proxy of actual contacts.

Consequently, one cannot easily chose between the 2 contradicting possibilities: people did not reduce their contacts, and the model is biased; or people did reduce their contacts, and the model is accurate. Not challenging models accuracy can lead to mistaking assumptions (“any difference between reality and model output is due to an intervention”) for conclusions (“the model shows the effect of the intervention”).

### 4.4 Policy Influence

Models have had large political influence in first implementing social distancing measures, and then justifying them.

The case of social distancing measures implementation is exemplified by the audition of the head of the French scientific council in the Senate, during hearings regarding the implementation of “health pass” in July 2021. It is stated that *“the model clearly shows that we are going to find ourselves in a complicated situation at the end of August”* Delfraissy [2021]. The models publicly released a few days later featured a range of possible outcomes from a low impact to a dire situation (see figure 4c). The French Conseil d’Etat then rejected legal challenges to this health pass implementation, citing these same models Conseil d’Etat [2021].

The second case of social distancing measures posterior justification is exemplified by Spain, when the first lockdown was judged unconstitutional by the Supreme Court, canceling all the fines that had been distributed. The Spanish president then justified the decision by stating that *“That is not me talking, but established by independent scientific studies - as a result of this lockdown, 450*,*000 lives have been saved”* Sanchez [2021]. While the source of the figure is not specified, it corresponds to the retrospective modelling study by Flaxman et al. [2020], which had already been cited by Spanish newspapers El Paìs [2020].

### 4.5 Ability to inform policy making

Some scenarios closely match reality, in most cases the best-case scenario. However these accurate scenarios are often part of reports displaying large uncertainty, often around or higher than 50% of the historical epidemic peak of spring 2020 (see section 3.2.3 and Figure 7). We insist on the fact that this uncertainty remains after hypotheses verification, and displays irreducible uncertainty.

For instance the July 2021 report features more than 100 scenarios regarding daily hospital admissions. After checking for vaccine uptake hypotheses, the uncertainty regarding epidemic peak remains substantive compared to the historical first wave peak of 3000 admissions. Scenarios span from 1000 to 2500 daily admissions in the best vaccine uptake configuration (uncertainty of 1500 admissions, or 50% of historical peak), and from 1500 to 4000 in the worst one (uncertainty of 2500, or 85% of historical peak). These ranges are entirely due to sheer uncertainties regarding modeled social distancing measures effectiveness, on which the policy-maker has no grasp.

### 4.6 Good practices for modelling scenario evaluation

In France, some modelers were also members of the scientific council designed to advise governments on policy-making. This might create a difficult situation where scientific advisers that help shape recommendations on non pharmaceutical interventions are both producers of epidemiological models used in hindsight to assess the effectiveness of the same interventions and evaluators of the epidemiological models accuracy. A good practice might be to have independent teams performing evaluation of epidemiological models.

Proper evaluation requires to systematically evaluate all of the published scenarios were comparison to reality is legitimate, to avoid cherry-picking of results in either direction. Publishing the scenarios data in open repositories would help such evaluations and their reproducibility.

## 5 Conclusion

In this systematic retrospective assessment of COVID scenarios in France, we find that the prospective scenarios proposed during the pandemic were generally more pessimistic than reality. Out of 10 reports, reality felt between the report best case and worst case scenarios in 3 report; in 6 reports, reality was below the best case scenario; in 1 report, reality was above the worst case scenario.

These results can inform expectations on how prospective scenarios informing public debate and policy making would relate to reality.

Systematic bias can emerge from systematically pessimistic assumptions in the model, or from a bias inherent to the model itself. If this latter case is true, using retrospective counterfactual scenarios to assess non pharmaceutical interventions effects may overestimate their efficacy.

Our work highlights the need for more systematic evaluation of the scenarios guiding policy makers in times of pandemic. It would be of interest to perform similar retrospective assessments in other countries, to see if our results are generalizable or specific to France.

## Supporting information

List of prospective scenarios

## Data Availability

All data produced are available online at https://github.com/maxime-langevin/retrospective_analysis

## Abbreviations

ICU: Intensive Care Units
COVID: Coronavirus Disease
MAE: Mean Absolute Error
MAPE: Mean Absolute Percentage Error
ME: Mean Error

## Code availability

Code supporting the results presented are available at the following github repository.

