## Supplementary material for "Retrospective analysis of Covid-19 hospitalization modelling scenarios which guided policy response in France"

### Supplementary Materials

#### Summary of the identified reports with prospective scenarios

Table S1: Identification of the 16 reports meeting criteria for retrospective comparison to reality. The 4 reports only reporting regional scenarios are not included in our analysis (Apr 12, Apr 28, May 12 and Nov 8, 2020), as well as the 2 reports only focusing on cases (Feb 21 and March 10, 2022), so our retrospective analysis includes a total of 10 reports.

| **Report date** | **Institution** | **National or Regional scale** | **Scenario endpoint** | | | **Identification through…** | | | |
|---|---|---|---|---|---|---|---|---|---|
| | | | **ICU** | **New Hosp** | **Cases** | **Scientific Council webpage** | **Pasteur website** | **EPIcx webpage** | **websearch** |
| Apr 12, 2020 | EPIcx | regional | x | | | x | | x | |
| Apr 28, 2020 | Pasteur | regional | x | | | | | | x |
| May 12, 2020 | EPIcx | regional | x | | | x | | x | |
| Oct 30, 2020 | Pasteur | National | x | | | | | | x |
| Nov 08, 2020 | EPIcx | regional | | x | | | | x | |
| Feb 08, 2021 | Pasteur | National | | x | | | x | | |
| Feb 14, 2021 | EPIcx | National + regional | | x | | | | x | |
| Feb 23, 2021 | Pasteur | National | | x | | x | x | | |
| Apr 26, 2021 | Pasteur | National | x | | | x | x | | |
| May 21, 2021 | Pasteur | National | x | x | | | x | | |
| Jul 26, 2021 | Pasteur | National | x | x | | | x | | |
| Aug 05, 2021 | Pasteur | National + regional | x | x | | | x | | |
| Oct 04, 2021 | Pasteur | National | | x | | x | x | | |
| Jan 07, 2022 | Pasteur | National | x | x | | x | x | | |
| Feb 21, 2022 | Pasteur | National | | | x | | | | |
| Mar 10, 2022 | Pasteur | National | | | x | | | | |

Table S2: Reports with prospective scenarios not meeting criteria for retrospective comparison to reality (see reason for exclusion below)

| Report date | Institution | Identification through… | | | |
|---|---|---|---|---|---|
| | | Scientific Council | Pasteur webpage | EPIcx webpage | websearch |
| Mar 14, 2020 | EPIcx | | | x | |
| Sept 25, 2020 | Pasteur | x | | | x |
| Sept 28, 2020 | EPIcx | x | | x | |
| Oct (?), 2020 | Pasteur | | | | x |
| Oct 26, 2020 | Pasteur | | x | | |
| Oct 26, 2020 | EPIcx | | | x | |
| Nov 17, 2020 | EPIcx | | | x | |
| Dec 3, 2020 | EPIcx | | | x | |
| Jan 12, 2021 | Pasteur | x | | | |
| Jan 16, 2021 | EPIcx | | | x | |
| Jan 24, 2021 | EPIcx | x | x | | |
| Jan 28, 2021 | EPIcx | x | x | | |
| Jan 28, 2021 | Pasteur | x | x | | |
| Feb 02, 2021 | EPIcx | | x | x | |
| Mar 11, 2021 | Pasteur | x | x | | |
| Jul 9, 2021 | Pasteur | | x | | |
| Jul 10, 2021 | EPIcx | | | x | |
| Nov 29, 2021 | Pasteur | x | x | | |
| Dec 2, 2021 | Pasteur | x | x | | |
| Dec 7, 2021 | Pasteur | | x | | |
| Dec 27, 2021 | Pasteur | x | x | | |

#### Excluded reports: reasons for exclusion

Table S3: exclusion reason for the  reports with prospective scenarios not included in our retrospective analysis.

| Date and Source | Institution | Reason for exclusion |
|---|---|---|
| March 14, 2020 Source (Report 8) | EPIcx | Only looks at the effect of school closure / telework, but the very same day of publication, national lockdown was announced, so hypotheses are not met anymore. Anyway, focuses only on 3 particular regions, but our retrospective is at the national scale |
| April 12, 2020 Source (Report 9) | EPIcx | Only at regional scale (our study reviews national scale scenarios) |
| April 28, 2020 Source | Pasteur | Only at regional scale (our study reviews national scale scenarios) |
| May 12, 2020 Source (Report 10) | EPIcx | Only at regional scale (our study reviews national scale scenarios) |
| September 25, 2020 Source | Pasteur | Starting September 26, localized restriction measures were announced, which were not accounted for in the report. Thus, the hypotheses of the report are no more valid.<br><br>Example of the restriction measures (source): bar and restaurant closures, sports halls closures, maximum number of people gathering limited, ban of large events… |
| September 28, 2020 Source (Report 16) | EPIcx | Only looks at teleworking effect but starting September 26, localized restriction measures were announced (bar and restaurant closures, sports halls closures, maximum number of people gathering limited, ban of large events), which were not accounted for in the report. see measures https://www.prefectures-regions.gouv.fr/ile-de-france/Region-et-institutions/L-action-de-l-Etat/Sante/COVID-19-le-point-sur-la-situation<br><br>Anyway, focuses only on 3 particular regions, but our retrospective is at the national scale |
| October (?), 2020 Source ? | Pasteur | We were not able to find the report, which to the best of our knowledge was not made publicly available. Indeed, according to franceinfo: |

| | | |
|---|---|---|
| | | *"The Minister of Health chose not to mention them [the models]. The concern to keep citizens on their toes has taken precedence over transparency, which would nevertheless require the scientific data to be made public, even if they are less catastrophic than what had been announced."* |
| October 26, 2020<br>Source | Pasteur | National lockdown was announced 2 days later, on October 28. Thus, the hypotheses are no more valid.<br><br>An update considering the lockdown effect was released on October 30, which is included in our analysis. |
| October 26, 2020<br>Source | EPIcx | "Lockdown is implemented starting week 45, 46, or 47, in each case lasting 4 weeks", but it was actually implemented on Friday October 30, on w44, 4 days after the publication of report. Anyway, the report only focuses on Ile-de-France region ("The current report focuses on Île-de-France only; analyses on other regions will follow."), and our retrospective is at the national scale |
| November 8, 2020<br>Source<br>(Report 21) | EPIcx | Only at regional scale (our study reviews national scale scenarios) |
| November 17, 2020<br>Source<br>(Report 23) | EPIcx | Only scenarios with lockdown exit on the 2/12 or the 21/12, while real exit from lockdown on the 15/12. But one might consider that reality should be between the 2/12 or the 21/12 scenarios. Anyway, the report only focuses on Ile-de-France region ("The report focuses on Île-de-France; analyses on other regions will follow"), and our analysis is only at national scale |
| December 3, 2020<br>Source<br>(Report 25) | EPIcx | Scenarios assume continued strict lockdown up to week 52. "lockdown projections after week 48 (Nov 23-29) assume the same conditions as in the first week lockdown and they do not envision the relaxation of restrictions starting November 28." So the scenarios do not take into account the easing of restrictions during weeks 48 to 52. |
| January 12, 2021<br>Source | Pasteur | Some preliminary projections regarding the impact of the alpha variant without further control are presented in the report, but remain very uncertain, both concerning the date of a potential rise in hospitalizations (from February to April) and the magnitude of the ICU peak (from 7 000 to 40 000).<br><br>The projections are clarified in subsequent reports (February 8, and February 23) that we analyze. |
| January 16, 2021<br>Source<br>(Report 26) | EPIcx | Does not account for the effect of the latest social distancing measure, enacted the same day as the publication date: "It does not account for the curfew anticipated to 6pm and extended to the national territory on January 16, 2021". For context, before that date, only about one fourth of the French departments were concerned by a 6 pm curfew, and the other three quarters departments were concerned by a 8 pm curfew. |
| January 24, 2021<br>Source<br>(Report 27) | EPIcx | See report in the annex of the Source<br><br>Does not account for the effect of the latest social distancing measure: "curfew of January 16 anticipated at 6pm and extended on the national territory is not considered here, as it is too early to estimate its effect". For context, before that date, only about one fourth of the French departments were concerned by a 6 pm curfew, and the other three quarters departments were concerned by a 8 pm curfew. |

| | | |
|---|---|---|
| | | Inclusion is debatable: extended curfew had been enacted 8 days ago, but the modelers still decided to publish these results, suggesting they considered the effect of the measure to be small<br><br>Anyway, the report is almost a duplicate, with assumptions and results very similar to reports 26, 27, 29. We only focus on the latest report of this period, i.e. Report 29 on Feb 14, 2021 |
| January 28, 2021<br>Source | Pasteur | See report in the annex of the Source<br><br>only presenting lockdown scenarios starting on February 1st of February 8th. Since no lockdown occurred until the end of March, we did not analyze this report. |
| January 28, 2021<br>Source<br>(Report 27 bis) | EPIcx | See report in the annex of the Source<br><br>Does not account for the effect of the latest social distancing measure: "La courbe grise correspond au scénario sans changement, c'est-à-dire sans interventions supplémentaires à ceux mises en place jusqu'au 10 janvier (couvre-feu à 18h00 dans 15 départements).". For context, before that date, only about one fourth of the French departments were concerned by a 6 pm curfew, and the other three quarters departments were concerned by a 8 pm curfew.<br><br>Inclusion is debatable: extended curfew had been enacted 12 days ago, but the modelers still decided to publish these results, suggesting they considered the effect of the measure to be small<br><br>Anyway, the report is almost a duplicate, with assumptions and results very similar to reports 26, 27, 29. We only focus on the latest report of this period, i.e. Report 29 on Feb 14, 2021 |
| February 2, 2021<br>Source<br>(Report 28) | EPIcx | Inclusion is debatable, as it is less clear than reports 26 and 27 on whether assumptions account for the anticipated curfew (seems to be accounted for): "Différentes évolutions de la trajectoire épidémique dans les semaines à venir sont aussi possibles, à cause de cette incertitude ou d'autre facteurs qui ne peuvent pas être anticipées (comme par exemple des comportements adaptatifs des individus au couvre-feu renforcé). Pour cette raison, on propose aussi 2 autres scenarii, avec Reff(non-VOC)=0.9 et 1.1".<br><br>Anyway, the report is almost a duplicate, with assumptions and results very similar to reports 26, 27, 29. We only focus on the latest report of this period, i.e. Report 29. |
| March 11, 2021<br>Source | Pasteur | All the figures are already presented in the Feb 23, 2021 report, which is already assessed in our retrospective. |
| July 9, 2021<br>Source | Pasteur | The implementation of a Health Pass was announced 3 days later, on July 12. Thus, the hypotheses made in the report are no longer valid.<br><br>Scenarios accounting for these new measures were published on July 26 and August 5, and are included in our analysis. |
| July 10, | EPIcx | |

| | | |
|---|---|---|
| 2020<br>Source | | |
| November 29, 2021<br>Source | Pasteur | The unexpected emergence of the Omicron variant of concern occurred a few days later, representing 15% of contaminations by mid-December and 60% by the end of December (source).<br><br>A specific report accounting for the omicron variant was published on January 7, and is included in our analysis. |
| December 2, 2021<br>Source | Pasteur | The unexpected emergence of the Omicron variant of concern occurred a few days later, representing 15% of contaminations by mid-December and 60% by the end of December (source).<br><br>A specific report accounting for the omicron variant was published on January 7, and is included in our analysis. |
| December 7, 2021<br>Source | Pasteur | Results already presented on the November 29 and December 2 reports. Moreover, the same limitations concerning the unexpected emergence of the omicron variant apply here. |
| December 27, 2021<br>Source | Pasteur | At the moment of the publication, strong uncertainties remained concerning the impact of the Omicron variant, with scenarios spanning from a straight decrease of hospitalizations below 1000 to a sharp spike beyond 15 000 (figure 5). As stated by the report:<br><br>*"Given the significant uncertainties regarding the severity and transmission advantage of the Omicron variant over the Delta variant, it is not possible to accurately quantify the impact that the Omicron wave will have on the healthcare system."*<br><br>The uncertainties are greatly reduced in a subsequent report (January 7, 2022), that we analyze. |
| February 21, 2021<br>Source | Pasteur | The prospective scenarios only concern COVID cases, which are not included in our criteria analysis (we only focus on hospitalizations and intensive care units). |
| March 10, 2021<br>Source | Pasteur | The prospective scenarios only concern COVID cases, which are not included in our criteria analysis (we only focus on hospitalizations and intensive care units). |

#### Selected reports: detailed analysis

Table S4: Analysis and hypotheses justification for all 10 reports included in our retrospective analysis.

<table>
<tr><th>Date and Source</th><th>Scenarios hypotheses</th><th>Kept hypotheses</th><th>Extracted figures and curves for hospital admissions or ICU beds</th></tr>
<tr><td>October 30, 2020<br><br>Pasteur<br><br>source</td><td>Only different R values: from 0.7 to 1.2</td><td>All R values kept</td><td>ICU beds<br>All the lines of the figure were extracted</td></tr>
<tr><td>February 8, 2021<br><br>Pasteur<br><br>source</td><td>Different hypotheses of the scenarios:<br><br>R of the historical virus:<br>● $R_{eff}$(non-VOC) = 0.9<br>● $R_{eff}$ (non-VOC) = 0.95 (baseline)<br>● $R_{eff}$ (non-VOC) = 1<br><br>Increase in alpha variant transmissibility compared to historical:<br>● +50% (baseline)<br>● +40%<br>● +70%<br><br>General assumptions and limits:<br>● Vaccination: impacts hospitalization but barely infections at the national level<br>● Does consider the extended 6 p.m. curfew effect and the effect of January 29 measures<br>● Climate effect not considered<br>● Say their hypothesis about vaccine compliance is optimistic<br>● Winter holidays not considered</td><td>We keep all hypotheses regarding the different values of $R_{eff}$ (non-VOC) (0.85, 0.9, 1)<br><br>We keep all hypotheses regarding increased transmissibility (+40%, +50+, +70%). Indeed, many months later the issue was not settled by modelers (see April 26 and May 21 scenarios).<br><br>We stop our comparison on March 30$^{nd}$, the moment when the national lockdown is implemented and the comparison to the “no lockdown” scenarios is no more legitimate.</td><td>Hospital Admissions<br><br>Figure 2A<br>Purple lines not extracted (lockdown and/or no vaccination scenarios).<br>Both dashed and plain red curve (curfew until March 29 + vaccination or no vaccination scenario) are extracted, since it is stated in the report that “In practice, the impact of the current campaign is likely to be intermediate between the scenarios with and without vaccination presented in Figure 2”<br><br>Figure 3A, 3C<br>Not extracted. Green and purple lines are lockdown scenarios, red line is curfew until March 15 then no restrictions scenario<br><br>Figure 4A, 4C<br>Not extracted, only lockdown scenarios<br><br>Figure 5A<br>Not extracted, only lockdown scenarios<br><br>Figure 6A</td></tr>
</table>

| | | | |
|---|---|---|---|
| | | | All lines extracted until the peak (marking March 1st lockdown effect). The different lines represent sensitivity analysis regarding $R_{eff}$(non-VOC)<br><br>Figure 7A, 7C<br>We extract the C figure which allows longer comparison than the A figure (lockdown on March 1st vs February 15th for A graph). On the C figure we extract all lines (sensitivity analysis regarding Variant Of Concern 40%, 50%).<br><br>Figure 8A and 9A<br>Not extracted, only lockdown scenarios |
| February 14, 2021 (Report 29)<br><br>EPIcx<br><br>source | <u>Different hypotheses of the scenarios:</u><br>effective R of historical virus:<br>● 0.95 (0.94-0.96) best estimate at the time<br>● 1.05 (+10%, relaxation social distancing )<br>● 0.85 (-10%, strengthening social distancing)<br><br><u>General assumptions and limits:</u><br>● only difference of alpha variant with historical is transmissibility: severity not increased<br>● cross-immunity between strains<br>● Increase in alpha variant VOC (variant of concern) transmissibility compared to historical strain: +50% (+40% and +60% not shown)<br>● vaccination effect will not be visible until April<br>● fitted to hospital admissions up to Feb 7, 2021 (week 5), which is 3 weeks after the extension of the curfew (Jan 16, see more details in the description of excluded reports Jan 16 or Feb 2). | We keep the 3 hypotheses regarding R of historical virus<br><br>We stop our comparison on March 22nd, the moment when the national lockdown is implemented and the comparison to the “no lockdown” scenarios is no more legitimate. | <u>Hospital Admissions</u><br>Figure 1: the gray curves from the 3 top panes (bottom panes are only for the Ile-de-France region).<br><br>Figure S: not extracted, as it is a sensitivity analysis of vaccination uptake, which only marginally changes the results in the short to medium term, as said by the modelers in the report |

| | | | |
|---|---|---|---|
| | • reduced susceptibility adolescents and children, reduced transmissibility children | | |
| February 23, 2021<br><br>Pasteur<br><br>source | <u>Different hypotheses of the scenarios:</u><br>• Historical virus R (0.94, 1.11, 1.19, 1.28)<br>• Change in historical virus R starting March 8th or February 22nd<br>• Variant Of Concern increased transmissibility (+50%, +60%, +70%)<br>• Vaccine efficacy on transmission: 0% vs 30% (but does not change the results during our study period, only later)<br>• Vaccine coverage: 70% in every age group (baseline) vs 90% for >65y<br><br><u>General assumptions and limits:</u><br>• Lockdown implemented March 22nd: we compare the result until this date<br>• Different rates of vaccine doses supplied per day, changing on April 1st: thus, does not change on our study period<br>• Vaccine efficacy on severe cases: 90%<br>• No effect of the B.1.1.7 on hospitalizations? | We keep all the hypotheses regarding the historical virus different R.<br><br>We keep all hypotheses regarding increased transmissibility (+50%, +60+ and +70%). Indeed, many months later the issue was not settled by modelers (see April 26 and May 21 scenarios).<br><br>For the hypothesis regarding vaccine efficacy and supply, they do not affect the hospitalization curves during our study period (see figure 4, 5G and 5H).<br><br>We stop our comparison on March 22nd, the moment when the national lockdown is implemented and the comparison to the "no lockdown" scenarios is no more legitimate. | <u>Hospital Admissions</u><br><br>Figure 2C<br>Plain red line extracted (vaccination scenario), dotted line not extracted (no vaccination scenario).<br><br>Figure 3<br>Not extracted, same as 2C for March with different lockdown scenarios starting March 22 (not our study period).<br><br>Figure 4A, 4B, 4C<br>Not extracted. Different lockdown strategies coupled with vaccination hypothesis, which does not change results before March 22nd, presented in figure 2C.<br><br>Figure 5B, 5D, 5F, 5H<br>Not extracted. Same as figures 5A, 5C, 5E, 5G but with stronger measures starting March 22, does not change our results for our study period.<br><br>Figures 5A, 5C, 5E, 5G<br>A: extracted, hypothesis regarding historic virus R (0.94, 1.11, 1.19, 1.28)<br>C: extracted, VOC transmissibility compared to historic variant (+50%, +60%, +70%)<br>E: extracted, same as A but change occurs on February 22 instead of March 8<br>G: not extracted, variations of vaccine efficacy but does not affect the results of 5A-5E figures on our study period |
| April 26, 2021<br><br>Pasteur<br><br>source | <u>Different hypotheses of the scenarios:</u><br>• VOC increased transmissibility (+60%, +40%)<br>• Hospitalization decrease more or less intense | We keep all hypotheses regarding VOC increased transmissibility (+60% and +40%)<br><br>We keep both hypotheses regarding hospitalization decrease rates | <u>Hospital Admissions</u><br><br>Figure 2<br>Not extracted since all scenarios presented here are also included in figure 3 |

| | | | |
|---|---|---|---|
| | ● Vaccine doses per day: 350k or 500k<br>● Different R for the non-VOC virus, reflecting the progressive end of restriction measures (1, 1.1, 1.2 and 1.3)<br><br>General assumptions and limits:<br>● B.1.1.7 (alpha) variant: +64% probability of hospitalization<br>● Vaccine efficacy: 90% on severe cases, 80% on infections, starting 2 weeks after dose 1<br>● Vaccine compliance: 85% for >65y, 70% for 18-64y<br>● Try to account for climate effect but still uncertainties<br>● Say their vaccine compliance hypothesis is optimistic<br>● Infected people have total immunity<br>● Does not consider the end of travel restrictions, nor re-opening of middle and high schools. | For vaccine doses per day, the actual pace was 500k, so we exclude the 350k scenarios (source)<br><br>We exclude the more pessimistic R=1.3, corresponding to a transmission comparable to summer 2020, when there were almost no restrictions | Figure 3A and 3C<br>Not extracted because did not match the vaccination doses hypothesis (350k vs 500-600k in reality)<br><br>Figure 3B and 3D<br>-Colors corresponding to different R. As said on the left, we exclude the R=1.3 (red). We extract all the other ones (orange, purple, blue).<br>-Dashed vs plain lines correspond to faster or slower hospitalization decrease. We extract both<br>-B vs D: different hypothesis corresponding to B.1.1.7 increased transmissibility (60% vs 40%). We extract both. |
| May 21, 2021<br><br>Pasteur<br><br>source | Different hypotheses of the scenarios:<br>● VOC increased transmissibility (+60%, +40%)<br>● Vaccine doses per day: 500k or 700k<br>● Different R for the non-VOC virus, reflecting the progressive end of restriction measures (1, 1.1, 1.2 and 1.3)<br>● Dynamic changes on June 9$^{th}$ or May 19$^{th}$ (reflecting released restrictions)<br><br>General assumptions and limits:<br>● B.1.1.7 variant: +64% probability of hospitalization | Delta variant:<br>After the publication there was the unexpected emergence of the delta variant, not considered in the scenarios. Its share in the total contaminations was 5% on June 7, 15% on June 21, and 50% on July 21 (source).<br>We stop our comparison at mid-June when the delta variant impact on ICU was still negligible.<br><br>We keep all hypothesis regarding VOC increased transmissibility | Hospital Admissions<br><br>Figure 1B, 1F, 1D, 1H<br>Not extracted because of unmet vaccination distribution hypotheses (700k/day)<br><br>Figures 1E, 1G<br>Changing dynamic on June 9$^{th}$ coupled with increased transmissibility of alpha variant of 60% or 40%. We extract both<br>The different R values (colors) are not affected on our study period<br><br>Figures 1A, 1C |

| | | | |
|---|---|---|---|
| | ● Vaccine efficacy: 90% on severe cases, 80% on infections, starting 2 weeks after dose 1<br>● Vaccine compliance: 85% for >65y, 70% for 18-64y<br>● Try to account for climate effect but still uncertainties<br>● Infected people have total immunity | For vaccine doses per day, the actual pace was 500k-600k, so we exclude the 700k scenarios (source) | Changing dynamic on May 19th coupled with increased transmissibility of alpha variant of 60% or 40% and different R values.<br>We extract all the curves.<br><br><br>ICU beds<br><br>Figure 3B, 3F, 3D, 3H<br>Not extracted because of unmet vaccination distribution hypotheses (700k/day)<br><br>Figures 3E, 3G<br>Changing dynamic on June 9th coupled with increased transmissibility of alpha variant of 60% or 40%. We extract both<br>The different R values (colors) are not affected on our study period<br><br>Figures 3A, 3C<br>Changing dynamic on May 19th coupled with increased transmissibility of alpha variant of 60% or 40% and different R values.<br>We extract all the curves. |
| July 26, 2021<br><br>Pasteur<br><br>source | Different hypotheses of the scenarios<br>● Vaccine doses per day: 500k, 700k or 800k<br>● Vaccine compliance for >60y (90% or 95%), 18-60y (70% or 90%), 12-17y (30%, 50% or 70%)<br>● Reduction of R due to NPIs (health pass, masks…): 0%, 10% or 25% (2, 1.8 or 1.5)<br>● Time spent in intensive care units: 14.6 days or 10 days<br><br>General assumptions and limits:<br>● Uncertainties regarding climate effect | We keep all scenarios regarding R values<br><br>We keep all scenarios regarding time spend in ICU<br><br>For the first week of the scenarios, actual vaccine supply was 600-700k/day. Until August 9 it was 500k-600k/day, then below 500k/day (source)<br>We thus keep the 500k/day and 700k per day scenarios<br><br>On October 1st, vaccine compliance was (source): | Hospital Admissions<br><br>Figure 3<br>Owing to vaccine compliance hypotheses, we focus on the bottom pane, 3rd from the left<br>We extract all the colors (hypotheses regarding R)<br>We extract plain and dashed lines (500k and 700k vaccines doses per day).<br><br><br>ICU beds<br><br>Figure 5<br>Owing to vaccine compliance hypotheses, we focus on the bottom pane, 3rd from the left<br>We extract all the colors (hypotheses regarding R) |

<table>
<tr><td></td><td><ul><li>Does not consider reduced vaccine efficacy with delta</li><li>Does not consider increased risk of hospitalization with delta compared to alpha variant (here for both variant risk of hospitalization is +64% compared to historical strain)</li><li>Vaccine efficacy: 2 weeks after dose 1, 90% against hospitalization, 80% against infection, 50% against transmission</li></ul></td><td><ul><li>90% for >65y</li><li>90% for 18-60y</li><li>We did not find the % for 12-17y, but it was 25% for 0-17y, at a time when vaccination was not possible for 0-12y, suggesting 50% of 70% for 12-17y. We keep the 70% scenario (in any case, for high vaccination rates in adults, scenarios are not affected by adolescents' vaccination. Furthermore, mistakenly not taking into account the 50% scenario would play in favor of the scenarios accuracy considering scenarios from this report overestimated reality)</li></ul></td><td>We extract plain and dashed lines (500k and 700k vaccines doses per day).<br><br>Figure 6<br>Same as figure 5 but for an ICU duration of 10 days. We extract the same curves.</td></tr>
<tr><td>August 5, 2021<br><br>Pasteur<br><br>source</td><td><u>Different hypotheses of the scenarios</u><ul><li>Reduction of R due to NPIs (health pass, masks…): 10%, 25% or 40%</li><li>Time spent in intensive care units: 10, 14 or 17 days</li><li>Vaccine doses per day: 600k, 700k or 800k</li></ul><br><u>General assumptions and limits:</u><ul><li>Vaccine compliance for >60y is 95%, 90% for 18-60y, and 30% for 12-17y</li><li>Does not consider decreased vaccine efficacy against delta => scenarios may be too optimistic</li><li>Does not consider increased risk of hospitalization with delta compared to alpha variant (here for both variant risk of hospitalization is +64% compared to historical strain</li></ul></td><td>We keep all scenarios regarding R values<br><br>We keep all scenarios regarding time spend in ICU<br><br>For the first week of the scenario, vaccine supply was 500k-600k/day, then below 500k/day (source)<br>We thus keep the 600k/day scenario, which is the closest one to actual figures</td><td><u>Hospital Admissions</u><br><br>Figure A<br>We only extract the red curve (600k/day vaccine doses) and not the blue and green curves (700 and 800 k/day).<br><br>Figure C<br>We extract all the curves (correspond to different R reduction values due to sanitary measures).<br><br><u>ICU beds</u><br><br>Figure F<br>We extract all curves</td></tr>
</table>

| | | | |
|---|---|---|---|
| | • Uncertainties regarding climate effect<br>• Once recovered infected people are totally immunized, "This may lead to overly optimistic projections"<br>• Vaccine efficacy: 2 weeks after dose 1, 90% against hospitalization, 80% against infection, 50% against transmission | | |
| October 4th, 2021<br><br>Pasteur<br><br>source | Different hypotheses of the scenarios:<br>• Winter climate impact: +33% (baseline), +20%, 40%<br>• Easing of restrictions and/or behaviors: same transmission as in June-July (strong easing); "current" (-40% compared to June-July) ; intermediate (-20% compared to June-July). Not possible to precisely account for the health pass effect but these scenarios try to account for it.<br>• Vaccine efficacy: 95% against hospitalizations, 60% against infection. Also, scenarios 90%-60% and 95%-80%<br><br>General assumptions and limits:<br>• Vaccine compliance by December 80% in adolescents and 90% in adults<br>• Vaccine efficacy: 50% against transmission given infected<br>• Delta variant increases hospitalization risk by 50% compared to alpha variant. | Omicron share in infections was ~1% on December 6th and ~15% on December 20th (source), so we stop our comparison at mid-December<br><br>We keep all the hypotheses regarding winter climate impact<br><br>We keep all the hypotheses regarding easing of restrictions and/or behaviors<br><br>We keep all the hypotheses regarding vaccine efficacy, since vaccine efficacy against delta variant of 60%-80% against infection and 90-95% are consistent with UKHSA consensus estimates (here and here) | Hospital Admissions<br><br>Figure 5 right<br>Not extracted, since already included in more complete figure 7 and 9<br><br>Figure 7<br>We do not extract bottom and right figures, which do not induce a change in our study period (hypotheses regarding the changes in measures / behaviors on November 15th, December 15th, or January 15th).<br>We do not extract the top left graph (change in measures / behaviors on October 15th) since it is already included in more complete figure 9.<br><br>Figure 9<br>We extract all the curves, presenting different hypotheses regarding measures/behavior and climate effect<br>We extract the 3 graphs, presenting the different assumptions regarding vaccine efficacy |

| | | | |
|---|---|---|---|
| | ● Alpha variant increases hospitalization risk by 42% compared to historical train | | |
| January 7, 2022<br><br>Pasteur<br><br>source | Different hypotheses of the scenarios:<br>● Assumptions regarding vaccine efficacy detailed in Table S5 below<br>● Time spent in hospital: 3, 4 or 6 days<br>● Combination of omicron severity and transmissibility: half of historical strain and high transmissibility (scenario 1, "most probable"); as severe as historical strain and intermediate transmissibility (scenario 2, "less probable); as severe as historical strain and high transmissibility (scenario 3, "less probable)<br>● Reduction of $R_0$ due to restrictions and/or change in behaviors: 0%, -10%, -20%<br><br>General assumptions and limits:<br>● 800k vaccine booster doses per day, supply unaffected by winter holidays<br>● Once hospitalized, vaccinated and unvaccinated people have same probability of going to ICU => may lead to overestimation in scenarios with low vaccine efficacy<br>● Time spent in intensive care units: 14 days<br>● Vaccine booster compliance of 95% among adults, 4 months after initial doses | We exclude scenarios 2 and 3 which suppose that the Omicron variant has an identical severity compared to the historic strain. These were the 2 scenarios characterized as "less probable" by the report.<br><br>We keep the pessimistic assumptions regarding vaccine efficacy, since they are the closest to the "consensus vaccine effectiveness estimates" published by the UK Health Security Agency (source)<br><br>We keep all assumptions regarding time spent in hospital<br><br>We keep all assumptions regarding $R_0$ reduction | ICU beds<br><br>Figure 2, scenario 1, bottom panel,<br>Not extracted because it corresponds to the optimistic assumptions regarding vaccine efficacy<br><br>Figure 4, scenario 1, bottom panel<br>This corresponds to the pessimistic assumptions regarding vaccine efficacy<br>We extract all 3 curves (corresponding to different time spent in hospital)<br><br>Hospital admissions<br><br>Figure 2, scenario 1, top panel,<br>Not extracted because it corresponds to the optimistic assumptions regarding vaccine efficacy<br><br>Figure 4, scenario 1, top panel<br>This corresponds to the pessimistic assumptions regarding vaccine efficacy<br>We extract all 3 curves (corresponding to different time spent in hospital) |

| | |
|---|---|
| | • For children (5-11y), vaccination supply of 30k/day, with vaccine compliance of 30% |

#### Discussion: vaccine efficacy hypotheses made in the scenarios of January 7 and comparison to reality

The following 3 tables describe vaccine efficacy assumptions in Jan 7, 2022 report (Table S5) and the consensus vaccine efficacy estimates by UKHSA of Feb 3 (Table S6), available at the time of the Feb 15 self-assessment report. We also show later UKHSA estimates of June 16 (Table S7), supporting that initial estimates were relevant.

We estimate that UKHSA figures (Table S6 and S7) are closer to the pessimistic vaccine efficacy hypotheses (red figures in Table S5). On the contrary, in their retrospective assessment of Feb 15, the modelers focus on optimistic vaccine efficacy hypotheses (green ones in Table S5). This explains the discrepancy between their retrospective and ours (figures 9c and 9d in our main article).

We found no discussion related to the vaccine efficacy hypothesis in the modeler's self-assessment, which would help to explain their rationale for selecting the optimistic scenarios.

Table S5: Vaccine efficacy assumptions in the Jan 7, 2022 report

| | Variant | Protection against infection | | Protection against hospitalization | |
|---|---|---|---|---|---|
| | | <6 months | >6 months | <6 months | >6 months |
| Infected and unvaccinated | Delta | 85% | 60% | 90% | 85% |
| | Omicron | 35% | 15% | 80% | 50% |
| 2 doses | Delta | 80% | 50% | 95% | 85% |
| | Omicron | 55% OR 40% | 25% OR 10% | 90% | 70% |
| Booster dose OR infected and vaccinated | Delta | 95% | 85% | 95% | 95% |
| | Omicron | 85% OR 60% | 70% OR 40% | 95% | 90% |

*Green figures are the optimistic vaccine efficacy assumptions, red are the pessimistic ones, and black are common to both assumptions. In their own retrospective assessment of February 15th, modelers use the green ones. For our assessment, we use the red ones, based on UK's Health Security Agency data available at the time of modelers' self-assessment on February 3rd (Table S6 below), and the later "Consensus vaccine effectiveness estimates" from June 16th (Table S7 below).*

Table S6: Vaccine efficacy estimates by UKSH on Feb 3, 2022, at the time of the retrospective self-assessment by modelers (Feb 15, 2022)

Source, screenshot of p14. UK Health Security Agency, COVID-19 vaccine surveillance report, Week 5, 3 February 2022

**Table 2. Summary of evidence on vaccine effectiveness against different outcomes (a) Omicron (b) Delta (all vaccines combined)**

a)

| | **Dose 2** | | | **Dose 3** | | |
|---|---|---|---|---|---|---|
| | **0-3 months** | **4-6 months** | **6+ months** | **0-3 months** | **4-6 months** | **6+ months** |
| Infection | Insufficient data | Insufficient data | Insufficient data | Insufficient data | Insufficient data | Insufficient data |
| Symptomatic disease | 25-70% | 5-30% | 0-10% | 50-75% | 40-50% | Insufficient data |
| Hospitalisation | 65-85% | 55-65% | 30-35% | 80-95% | 75-85% | Insufficient data |
| Mortality | Insufficient data | Insufficient data | 40-70% | 85-99% | Insufficient data | Insufficient data |

b)

| | **Dose 2** | | | **Dose 3** | | |
|---|---|---|---|---|---|---|
| | **0-3 months** | **4-6 months** | **6+ months** | **0-3 months** | **4-6 months** | **6+ months** |
| Infection | 65-80% | 50-65% | Insufficient data | Insufficient data | Insufficient data | Insufficient data |
| Symptomatic disease | 65-90% | 45-65% | 40-60% | 90-99% | 90-95% | Insufficient data |
| Hospitalisation | 95-99% | 80-90% | 70-85% | 95-99% | Insufficient data | Insufficient data |
| Mortality | 95-99% | 90-95% | 80-99% | 95-99% | Insufficient data | Insufficient data |

| | |
|---|---|
| High Confidence | Evidence from multiple studies which is consistent and comprehensive |
| Medium Confidence | Evidence is emerging from a limited number of studies or with a moderately level of uncertainty |
| Low Confidence | Little evidence is available at present and results are inconclusive |

Table S7: Later consensus vaccine efficacy estimates by UKSH on June 16, 2022

*Source, screenshot of p13* UK Health Security Agency, COVID-19 vaccine surveillance report, Week 45, 16 June 2022

#### Consensus vaccine effectiveness estimates

Table 3 summarises consensus estimates of vaccine effectiveness against different outcomes that have been reached by the UK Vaccine Effectiveness Expert Panel. These take into account estimates from UK studies by public health agencies and academic groups as well as international data.

**Table 3. Consensus estimates of vaccine effectiveness against the Omicron variant**

| Vaccine product for primary course | Outcome | Second dose: 0 to 3 months | Second dose: 4 to 6 months | Second dose: 6+ months | Booster dose: All Periods | Booster dose: 0 to 3 months | Booster dose: 4 to 6 months | Booster dose: 6+ months |
|---|---|---|---|---|---|---|---|---|
| AstraZeneca | All Infection | 30% (20 to 40%) | 0 to 30% (range only) | 0% (0 to 10%) | See Individual Periods | 45% (35 to 55%) | 15% (0 to 30%) | 0% (0 to 10%) |
| | Symptomatic | 40% (30 to 50%) | 20% (5 to 30%) | 5% (0 to 5%) | See Individual Periods | 60% (50 to 70%) | 40% (30 to 50%) | 10% (0 to 20%) |
| | Hospitalisation | 85% (60 to 90%) | 70% (50 to 75%) | 65% (45 to 85%) | See Individual Periods | 90% (85 to 95%) | 85% (85 to 95%) | 70% (50 to 85%) |
| | Mortality | Insufficient Data | Insufficient Data | Insufficient Data | See Individual Periods | 90% (85 to 98%) | Insufficient Data | Insufficient Data |
| | Transmission | Insufficient Data | Insufficient Data | Insufficient Data | Insufficient Data | Insufficient Data | Insufficient Data | Insufficient Data |
| Moderna | All Infection | 30% (20 to 40%) | 0 to 30% (range only) | 30% (10 to 50%) | See Individual Periods | 45% (35 to 55%) | 15% (0 to 30%) | 0% (0 to 10%) |
| | Symptomatic | 55% (35 to 75%) | 30% (15 to 35%) | 15% (10 to 20%) | See Individual Periods | 65% (55 to 75%) | 40% (30 to 50%) | 10% (0 to 20%) |
| | Hospitalisation | 85 to 95% (range only) | 75 to 85% (range only) | 55 to 90% (range only) | See Individual Periods | 85 to 95% (range only) | Insufficient Data | Insufficient Data |
| | Mortality | Insufficient Data | Insufficient Data | Insufficient Data | Insufficient Data | Insufficient Data | Insufficient Data | Insufficient Data |
| | Transmission | Insufficient Data | Insufficient Data | Insufficient Data | Insufficient Data | Insufficient Data | Insufficient Data | Insufficient Data |
| Pfizer | All Infection | 30% (20 to 40%) | 0 to 30% (range only) | 20% (10 to 30%) | See Individual Periods | 45% (35 to 55%) | 15% (0 to 30%) | 0% (0 to 10%) |
| | Symptomatic | 50% (30 to 65%) | 20% (15 to 30%) | 15% (10 to 15%) | See Individual Periods | 65% (55 to 75%) | 45% (35 to 55%) | 10% (0 to 20%) |
| | Hospitalisation | 90% (85 to 95%) | 80% (75 to 85%) | 70% (55 to 90%) | See Individual Periods | 90% (85 to 95%) | 85% (85 to 95%) | 70% (50 to 85%) |
| | Mortality | Insufficient Data | Insufficient Data | Insufficient Data | See Individual Periods | 90% (85 to 98%) | Insufficient Data | Insufficient Data |
| | Transmission | Insufficient Data | Insufficient Data | Insufficient Data | 0 to 25% (range only) | Insufficient Data | Insufficient Data | Insufficient Data |

Booster data is based on use of the Moderna or Pfizer vaccines as a booster.
This table provides overall estimates but there may be variation by age group or other clinical or demographic factors.

| | |
|---|---|
| High Confidence | Evidence from multiple studies which is consistent and comprehensive |
| Medium Confidence | Evidence is emerging from a limited number of studies or with a moderately level of uncertainty |
| Low Confidence | Little evidence is available at present and results are inconclusive |

### Self-assessment by modelers

Figure S1: Scenarios publicly compared to reality by modelers (red) or not (gray) compared to reality (black line)

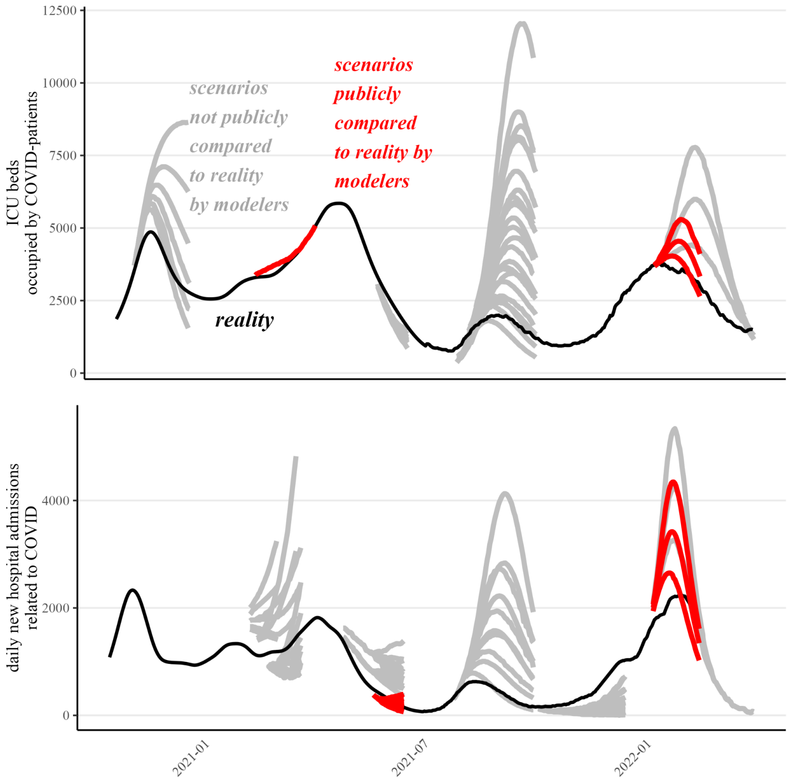
